# Can Auxiliary Indicators Improve COVID-19 Forecasting and Hotspot Prediction?

**DOI:** 10.1101/2021.06.22.21259346

**Authors:** Daniel J. McDonald, Jacob Bien, Alden Green, Addison J. Hu, Nat DeFries, Sangwon Hyun, Natalia L. Oliveira, James Sharpnack, Jingjing Tang, Robert Tibshirani, Valérie Ventura, Larry Wasserman, Ryan J. Tibshirani

**Author notes:** J.B., A.G., and A.J.H. contributed equally to this work.

## Abstract

Short-term forecasts of traditional streams from public health reporting (such as cases, hospitalizations, and deaths) are a key input to public health decision-making during a pandemic. Since early 2020, our research group has worked with data partners to collect, curate, and make publicly available numerous real-time COVID-19 indicators, providing multiple views of pandemic activity in the U.S. This paper studies the utility of five such indicators—derived from de-identified medical insurance claims, self-reported symptoms from online surveys, and COVID-related Google search activity—from a forecasting perspective. For each indicator, we ask whether its inclusion in an autoregressive (AR) model leads to improved predictive accuracy relative to the same model excluding it. Such an AR model, without external features, is already competitive with many top COVID-19 forecasting models in use today. Our analysis reveals that (a) inclusion of each of these five indicators improves on the overall predictive accuracy of the AR model; (b) predictive gains are in general most pronounced during times in which COVID cases are trending in “flat” or “down” directions; (c) one indicator, based on Google searches, seems to be particularly helpful during “up” trends.

---

Tracking and forecasting indicators from public health reporting streams—such as confirmed cases and deaths in the COVID-19 pandemic—is crucial for understanding disease spread, correctly formulating public policy responses, and rationally planning future public health resource needs. A companion paper [1] describes our research group’s efforts, beginning in April 2020, in curating and maintaining a database of real-time indicators that track COVID-19 activity and other relevant phenomena. The signals (a term we use synonomously with “indicators”) in this database are accessible through the COVIDcast API [2], as well as associated R [3] and Python [4] packages, for convenient data fetching and processing. In the current paper, we quantify the utility provided by a core set of these indicators for two fundamental prediction tasks: probabilistic forecasting of COVID-19 case rates and prediction of future COVID-19 case hotspots (defined by the event that a relative increase in COVID-19 cases exceeds a certain threshold).

At the outset, we should be clear that our intent in this paper is *not* to provide an authoritative take on cutting-edge COVID-19 forecasting methods. Similarly, some authors, e.g., [5], have pointed out numerous mishaps of forecasting during the pandemic, and it is not our general intent to fix them here. Instead, we start with a basic and yet reasonably effective predictive model for predicting future trends in COVID-19 cases, and present a rigorous, quantitative assessment of the added value provided by auxiliary indicators, that are derived from data sources that operate outside of traditional public health streams. In particular, we consider five indicators derived from de-identified medical insurance claims, self-reported symptoms from online surveys, and COVID-related Google searches.

To assess this value in as direct terms as possible, we base our study around a very simple basic model: an autoregressive model, in which COVID cases in the near future are predicted using a linear combination of COVID cases in the near past. Forecasting carries a rich literature, offering a wide range of sophisticated techniques, see, e.g., [6] for a review; however, we purposely avoid enhancements such as order selection, correction of outliers/anomalies in the data, and inclusion of regularization or nonlinearities. Similarly, we do not account for other factors that may well aid in forecasting, such as age-specific effects, holiday adjustments, and the effects of public health mandates. All that said, despite its simplicity, the basic autoregressive model that we consider in this paper exhibits competitive performance (see the Supplementary Materials for details) with many of the top COVID-19 case forecasters submitted to the U.S. COVID-19 Forecast Hub [7], which is the official source of forecasts used in public communications by the U.S. CDC. The strong performance of the autoregressive model here is in line with the fact that simple, robust models have also consistently been among the best-performing ones for COVID-19 death forecasting [8].

In the companion paper [1], we analyze correlations between various indicators and COVID case rates. These correlations are natural summaries of the contemporaneous association between an indicator and COVID cases, but they fall short of delivering a satisfactory answer to the question that motivates the current article: is the information contained in an indicator demonstrably useful for the prediction tasks we care about? Note that even lagged correlations cannot deliver a complete answer. Demonstrating *utility for prediction* is a much higher standard than simply asking about correlations; to be useful in forecast or hotspot models, an indicator must provide relevant information that is not otherwise contained in past values of the case rate series itself (cf. the pioneering work on Granger causality [9, 10], as well as the further references given below). We assess this directly by inspecting the difference in predictive performance of simple autoregressive models trained with and without access to past values of a particular indicator.

We find that each of the five indicators we consider— two based on COVID-related outpatient visits from medical insurance claims, one on self-reported symptoms from online surveys, and one on Google searches for anosmia or ageusia—provide an overall improvement in accuracy when incorporated into the autorgressive model. This is true both for COVID-19 case forecasting and hotspot prediction. Further analysis reveals that the gains in accuracy depend on the pandemic’s dynamics at prediction time: the biggest gains in accuracy appear during times in which cases are “flat” or trending “down”; but the indicator based on Google searches offers a most notable improvement when cases are trending “up”.

Careful handling of data revisions plays a key role in our analysis. Signals computed from surveillance streams are often subject to latency and/or revision. For example, a signal based on aggregated medical insurance claims may be available after just a few days, but it can then be substantially revised over the next several weeks as additional claims are submitted and/or processed late. Correlations between such a signal and case rates calculated “after the fact” (i.e., computed retrospectively, using the finalized values of this signal) will not deliver an honest answer to the question of whether this signal would have been useful in real time. Instead, we build predictive models using only the data that would have been available *as of* the prediction date, and compare the ensuing predictions in terms of accuracy. The necessity of real-time data for honest forecast evaluations has been recognized in econometrics for a long time, see [11–21]; but it is often overlooked in epidemic forecasting despite its critical importance, see, e.g., [22].

Finally, it is worth noting that examining the importance of additional features for prediction is a core question in inferential statistics and econometrics, with work dating back to at least [9]. Still today, drawing rigorous inference based on predictions, without (or with lean) assumptions, is an active field of research from both the applied and theoretical angles; see, e.g., [23–32]. Our take in the current work is in line with much of this literature; however, in order to avoid making any explicit assumptions, we do not attempt to make formal significance statements, and instead, broadly examine the stability of our conclusions with respect to numerous modes of analysis.

## 1 Methods

### 1.1 Signals and Locations

We consider prediction of future COVID-19 case rates or case hotspots (to be defined precisely shortly). By case rate, we mean the case count per 100,000 people (the standard in epidemiology). We use reported case data aggregated by JHU CSSE [33], which, like the auxiliary indicators that we use to supplement the basic autoregressive models, is accessible through the COVIDcast API [2].

The indicators we focus on provide information not generally available from standard public health reporting. Among the many auxiliary indicators collected in the API, we study the following five:

- Change Healthcare COVID-like illness (CHNG-CLI): The percentage of outpatient visits that are primarily about COVID-related symptoms, based on de-identified Change Healthcare claims data.
- Change Healthcare COVID (CHNG-COVID): The percentage of outpatient visits with confirmed COVID-19, based on the same claims data.
- COVID Trends and Impact Survey COVID-like illness in the community (CTIS-CLI-in-community): The estimated percentage of the population who know someone in their local community who is sick, based on Delphi’s COVID Trends and Impact Survey, in partnership with Facebook.
- Doctor Visits COVID-like illness (DV-CLI): The same as CHNG-CLI, but computed based on deidentified medical insurance claims from other health systems partners.
- Google search trends for anosmia and ageusia (Google-AA): A measure of Google search volume for queries that relate to anosmia or ageusia (loss of smell or taste), based on Google’s COVID-19 Search Trends data set.

We choose these indicators because, conceptually speaking, they measure aspects of an individual’s disease progression that would plausibly precede the occurence of (at worst, co-occur with) the report of a positive COVID-19 test, through standard public health reporting streams.

For more details on the five indicators (including how these are precisely computed from the underlying data streams) we refer to https://cmu-delphi.github.io/delphi-epidata/api/covidcast_signals.html, and the companion paper on the COVIDcast API and its signals [1]. For CTIS in particular, we refer to the companion paper [34]. For the Google COVID-19 Search Trends data set, see [35]; see also [36, 37] for a justification of the relevance of anosmia or ageusia to COVID infection.

As for geographic resolution, we consider the prediction of COVID-19 case rates and hotspots aggregated at the level of an individual *hospital referral region* (HRR). HRRs correspond to groups of counties in the United States within the same hospital referral system. The Dartmouth Atlas of Healthcare Policy [38], defines these 306 regions based on a number of characteristics. They are contiguous regions such that most of the hospital services for the underlying population are performed by hospitals within the region. Each HRR also contains at least one city where major procedures (cardiovascular or neurological) are performed. The smallest HRR has a population of about 125,000. While some are quite large (such as the one containing Los Angeles, which has more than 10 million people), generally HRRs are much more homogenous in size than the (approximately) 3200 U.S. counties, and they serve as a nice middle ground in between counties and states.

HRRs, by their definition, would be most relevant for forecasting hospital demand. We have chosen to focus on cases (forecasting and predicting hotspots) at the HRR level because the indicators considered should be more useful in predicting case activity rather than hospital demand, as the former is intuitively more contemporaneous to the events that are measured by the given five indicators. Predicting case rates (and hotspots) at the HRR level is still a reasonable goal in its own right; and moreover, it could be used to feed predicted case information into downstream hospitalization models.

### 1.2 Vintage Training Data

In this paper, all models are fit with “vintage” training data. This means that for a given prediction date, say, September 28, 2020, we train models using data that would have been available to us *as of* September 28 (imagine that we can “rewind” the clock to September 28 and query the COVIDcast API to get the latest data it would have had available at that point in time.) This is possible because of the COVIDcast API’s comprehensive data versioning system (described in more detail in the companion paper [1]). We also use the evalcast R package [39], which streamlines the process of training arbitrary prediction models over a sequence of prediction dates, by constructing the proper sequence of vintage training data sets.

Vintage training data means different things, in practice, for different signals. The three signals based on medical claims, CHNG-CLI, CHNG-COVID, and DV-CLI, are typically 3-5 days latent, and subject to a considerable but regular degree of revision or “backfill” after their initial publication date. The survey-based signal, CTIS-CLI-in-community, is 2 days latent, and rarely undergoes any revision at all. The target variable itself, reported COVID-19 case rates, is 1 day latent, and exhibits frequent, unpredictable revisions after intial publication. Compared to the pattern of revisions in the medical claims signals, which are much more systematic in nature, revisions in case reports can be highly erratic. Big spikes or other anomalies can occur in the data as reporting backlogs are cleared, changes in case definitions are made, etc. Groups like JHU CSSE then work tirelessly to correct such anomalies after first publication (e.g., they will attempt to back-distribute a spike when a reporting backlog is cleared, by working with a local authority to figure out how this should best be done), which can result in very nontrivial revisions. See Figure 1 for an example.

**Figure 1:**
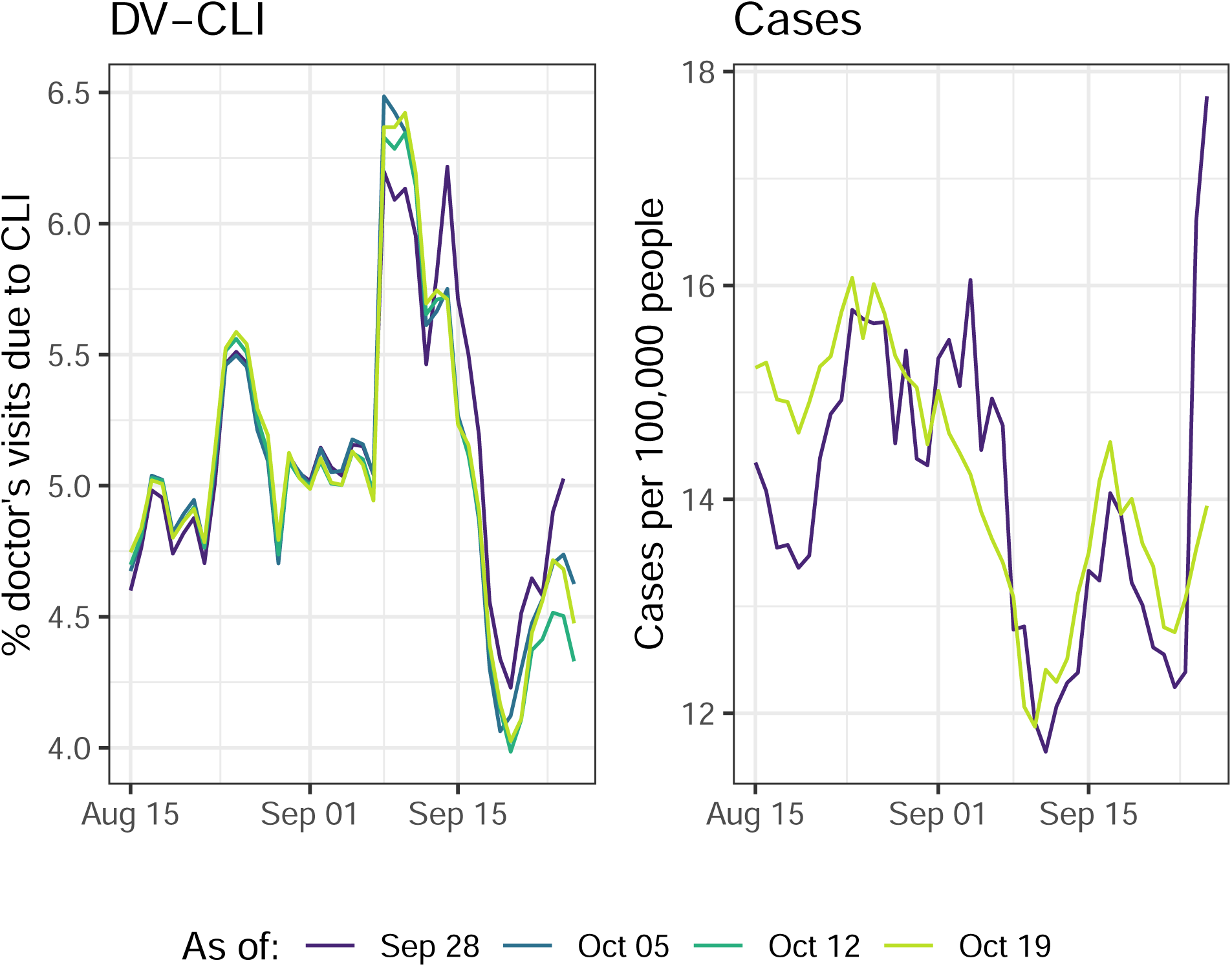
Revision behavior for two indicators in the HRR containing Charlotte, North Carolina. Each colored line corresponds to the data as reported on a particular date (*as of* dates varying from September 28 through October 19). The left panel shows the DV-CLI signal, which was regularly revised throughout the period, though the effects fade as we look further back in time. In contrast, the right panel shows case rates reported by JHU CSSE (smoothed with a 7-day trailing average), which remain “as reported” on September 28, with a spike towards the end of this period, until a major correction is made on October 19, which brings this down and affects all prior data as well.

Lastly, our treatment of the Google-AA signal is different from the rest. Because Google’s team did not start publishing this signal until early September, 2020, we do not have true vintage data before then; the latency of the signal was always at least one week through 2020. However, unlike the claims-based signals, there is no reason for revisions to occur after initial publication, and furthermore, the latency of the signal is not an unavoidable property of the data type, so we simply use finalized signal values, with zero latency, in our analysis.

### 1.3 Analysis Tasks

To fix notation, let *Y*_*ℓ,t*_ denote the 7-day trailing average of COVID-19 case incidence rates in location (HRR) *ℓ* and at time (day) *t*. To be clear, this is the number of new daily reported cases per 100,000 people, averaged over the 7-day period *t*−6, …, *t*. The first task we consider—*forecasting* —is to predict *Y*_*ℓ,t*+*a*_ for each “ahead” value *a* = 7, …, 21. The second task—*hotspot prediction*—is to predict a binary variable defined in terms of the relative change of *Y*_*ℓ,t*+*a*_ (relative to its value one week prior, *Y*_*ℓ,t*+*a*−7_), again for each *a* = 7, …, 21.

Why do we define the response variables via 7-day averaging? The short answer is robustness: averaging stabilizes the case time series, and moderates uninteresting artifacts like day-of-the-week effects in the data. We note that we can also equivalently view this (equivalent up to a constant factor) as predicting the HRR-level case incidence rate *summed* over some 7-day period in the future, and predicting a binary variable derived from this.

In what follows, we cover more details on our two analysis tasks. Table 1 presents a summary.

**Table 1:** Summary of forecasting and hotspot prediction tasks considered in this paper.

|  | Forecasting | Hotspot prediction |
| --- | --- | --- |
| Response variable | $Y_{\ell,t}$ (7-day trailing average of COVID-19 case incidence rates, per location $\ell$ and time $t$) | $Z_{\ell,t} = \mathbf{1}(Y_{\ell,t} \geq 1.25 \cdot Y_{\ell,t-7})$ (indicator that $Y_{\ell,t}$ grows by more than 25% relative to the preceding week) |
| Geographic resolution | Hospital referral region (HRR) | Hospital referral region (HRR) |
| Forecast period | June 9–December 31, 2020 | June 16–December 31, 2020 |
| Model type | Quantile regression | Logistic regression |
| Evaluation metric | Weighted interval score (WIS) | Area under curve (AUC) |

#### Dynamic Retraining

For each prediction date *t*, we use a 21-day trailing window of data to train our forecast or hotspot prediction models (so, e.g., the trained models will differ from those at prediction date *t* − 1). This is done to account for (potential) nonstationarity. For simplicity, the forecasting and hotspot prediction models are always trained on data across all HRRs (i.e., the coefficients in the models do not account for location-specific effects).

#### Prediction Period

In our analysis, we let the prediction date *t* run over each day in between early/mid June and December 31, 2020. The precise start date differs for forecasting and hotspots prediction; for each task it was chosen to be the earliest date at which the data needed to train all models was available, which ends up being (per our setup, with 21 days of training data and lagged values of signals for features, as we will detail shortly) June 9, 2020 for forecasting, and June 16, 2020 for hotspot prediction. (The bottleneck here is the CTIS-CLI-in-community signal, which does not exist before early April 2020, when the survey was first launched).

#### Forecasting Models

Recall *Y*_*ℓ,t*_ denotes the 7-day trailing average of COVID-19 case incidence rates in location *ℓ* and at time *t*. Separately for each *a* = 7, …, 21, to predict *Y*_*ℓ,t*+*a*_ for ahead value *a*, we consider a simple probabilistic forecasting model of the form:

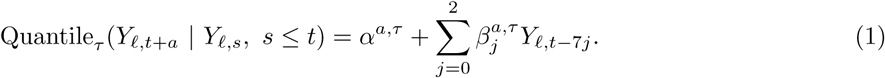

This model uses current case rates, and the case rates 7 and 14 days ago, in order to predict (the quantiles of) case rates in the future. We consider a total of 7 quantile levels (chosen in accordance with the county-level quantile levels suggested by the COVID-19 Forecast Hub),

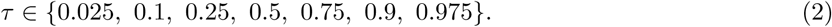

We fit (1) using *quantile regression* [40–42] separately for each *τ*, using data from all 306 HRRs, and within each HRR, using the most recent 21 days of training data. This gives us 6,426 training samples for each quantile regression problem.

In addition to this pure autoregressive model, we also consider five probabilistic forecasting models of the form:

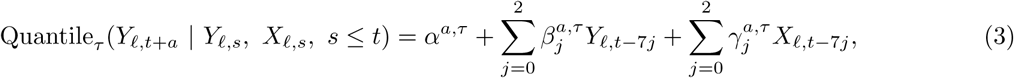

where *X*_*ℓ,t*_ denotes any one of the five auxiliary indicators—CHNG-CLI, CHNG-COVID, CTIS-CLI-in-community, DV-CLI, or Google-AA—at location *ℓ* and time *t*. Note that we apply the same lags (current value, along with the values 7 and 14 days ago) for the auxiliary indicators as we do for the case rates. Training then proceeds just as before: we use the same 7 quantile levels in (2), and fit quantile regression separately for each level *τ*, using data from all 306 HRRs and a trailing window of 21 days of training data. At prediction time, in order to avoid crossing violations (that is, for two levels *τ* ^′^ *> τ*, the predicted quantile at level *τ* exceeds the predicted quantile at level *τ* ^′^), we apply a simple post hoc sorting. See Figure 2 for an example forecast.

**Figure 2:**
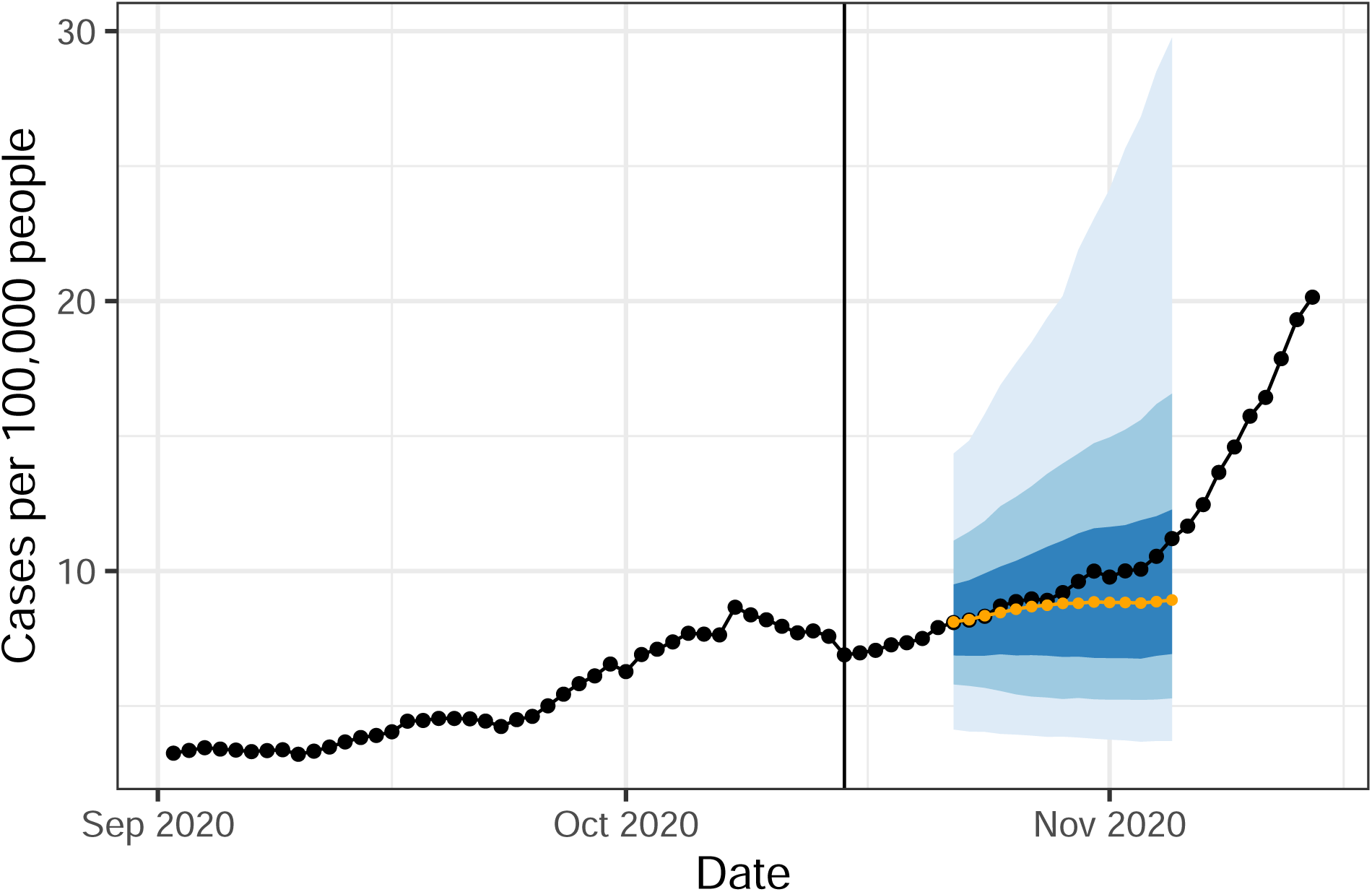
Forecast for the HRR containing New York City from an autoregressive model made on October 15 (vertical line). The fan displays 50%, 80% and 95% intervals while the orange curve shows the median forecast. The black curve shows “finalized” data, as reported in May 2021.

#### Hotspot Prediction Models

Define the binary indicator:

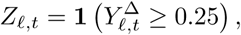

where we use the notation 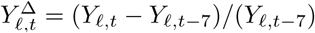. In other words, *Z*_*ℓ,t*_ = 1 if the number of newly reported cases over the past 7 days has increased by at least 25% compared to the preceding week. When this occurs, we say location *ℓ* is a *hotspot* at time *t*. Empirically, this rule labels about 27% of location-time pairs as hotspots, during the prediction period (June 16–December 31, 2020).

We treat hotspot prediction as a binary classification problem and use a setup altogether quite similar to the forecasting setup described previously. Separately for each *a* = 7, …, 21, to predict *Z*_*ℓ,t*+*a*_, we consider a simple logistic model:

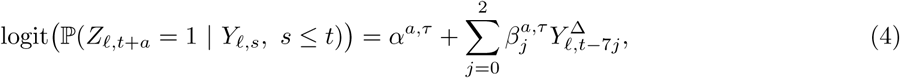

where logit(*p*) = log(*p/*(1 − *p*)), the log-odds of *p*.

In addition to this pure autoregressive model, we also consider five logistic models of the form:

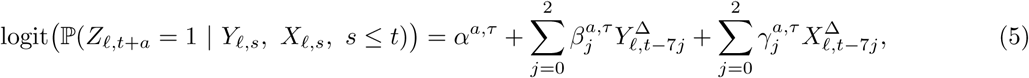

where we use 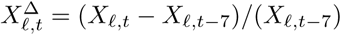, and again *X*_*ℓ,t*_ stands for any of the five auxiliary indicators at location *ℓ* and time *t*. We fit the above models, (4), (5), using logistic regression, pooling all 306 HRRs and using a 21-day trailing window for the training data.

An important detail is that in hotspot prediction we remove all data from training and evaluation where, on average, fewer than 30 cases (this refers to a count, not a rate) are observed over the prior 7 days. This avoids having to make arbitrary calls for a hotspot (or lack thereof) based on small counts.

### 1.4 Evaluation Metrics

For forecasting, we evaluate the probabilistic forecasts produced by the quantile models in (1) and (3) using *weighted interval score* (WIS), a quantile-based scoring rule; see, e.g., [43]. WIS is a proper score, which means that its expectation is minimized by the population quantiles of the target variable. The use of WIS in COVID-19 forecast scoring was proposed by [44]; WIS is also the main evaluation metric used in the COVID-19 Forecast Hub. More broadly, the specific form of WIS used here is a standard metric in the forecasting community for evaluating quantile-based probabilistic forecasts, just as mean squared forecast error is standard for point forecasts.

WIS is typically defined for quantile-based forecasts where the quantile levels are symmetric around 0.5. This is the case for our choice in (2). Let *F* be a forecaster comprised of predicted quantiles *q*_*τ*_ parametrized by a quantile level *τ*. In the case of symmetric quantile levels, this is equivalent to a collection of central prediction intervals (*f*_*α*_, *u*_*α*_), parametrized by an exclusion probability *α*. The WIS of the forecaster *F*, evaluated at the target variable *Y*, is defined by:

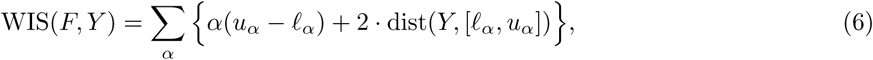

where dist(*a, S*) is the distance between a point *a* and set *S* (the smallest distance between *a* and an element of *S*). Note that, corresponding to (2), the exclusion probabilities are *α* ∈ {0.05, 0.2, 0.5, 1}, resulting in 4 terms in the above sum. By straightforward algebra, it is not hard to see WIS has an alternative representation in terms of the predicted quantiles themselves:

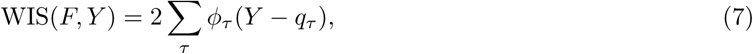

where *ϕ*_*τ*_ (*x*) = *τ* |*x*| for *x* ≥ 0 and *ϕ*_*τ*_ (*x*) = (1−*τ*) |*x*| for *x <* 0, which is often called the “tilted absolute” loss. While (7) is more general (it can accomodate asymmetric quantile levels), the first form in (6) is typically preferred in presentation, as the score nicely decouples into a “sharpness” component (first term in each summand) and an “under/overprediction” component (second term in each summand). But the second form given in (7) is especially noteworthy in our current study because it reveals WIS to be the same as the quantile regression loss that we use to train our forecasting models.

For hotspot prediction, we evaluate the probabilistic classifiers produced by the logistic models in (4) and (5) using the area under the curve (AUC) of their true positive versus false positive rate curve (which is traced out by varying the discrimination threshold).

The primary aggregation scheme that we will use in model evaluation and comparisons will be to average WIS per forecaster at ahead value *a* over all forecast dates *t* and locations *ℓ*; and similarly, to compute AUC per classifier at ahead value *a* over all forecast dates *t* and locations *ℓ*.

### 1.5 Other Considerations

#### Missing Data Imputation

Over the prediction period, all auxiliary indicators are available (in the proper vintage sense) for all locations and prediction times, except for the Google-AA signal, which is only observed for an average of 105 (of 306) HRRs. Such missingness occurs because the COVID-19 search trends data is constructed using differential privacy methods [45], and a missing signal value means that the level of noise added in the differential privacy mechanism is high compared to the underyling search count. In other words, values of the Google-AA signal are clearly *not* missing at random. It seems most appropriate to impute missing values by zero, and this is what we do in our analysis.

#### Backfill and Nowcasting

As described previously, the auxiliary indicators defined in terms of medical claims (CHNG-CLI, CHNG-COVID, and DV-CLI) undergo a significant and systematic pattern of revision, or “backfill”, after their initial publication. Given their somewhat statistically-regular backfill profiles, it would be reasonable to attempt to estimate their finalized values based on vintage data—a problem we refer to as *nowcasting*—as a pre-processing step before using them as features in the models in (3) and (5). Nowcasting is itself a highly nontrivial modeling problem, and we do not attempt it in this paper (it is a topic of ongoing work in our research group), but we note that nowcasting would likely improve the performance of the models involving claims-based signals in particular.

#### Spatial Heterogeneity

Some signals have a significant amount of spatial heterogeneity, by which we mean their values across different geographic locations are not comparable. This is the case for the Google-AA signal (due to the way in which the Google search trends time series is self-normalized, see [35]) and the claims-based signals (due to market-share differences, and/or differences in health-seeking behavior). Such spatial heterogeneity likely hurts the performance of the predictive models that rely on these signals, because we train the models on data pooled over all locations. In the current paper, we do not attempt to address this issue, and we simply note that location-specific effects (or pre-processing to remove spatial bias) would likely improve the performance of the models involving Google-AA and the claims-based indicators.

## 2 Results

Here, and in what follows, we will use “AR” to refer to the pure autoregressive model both in forecasting, (1), and in hotspot prediction, (4) (the reference to the prediction task should always be clear from the context). We will also use the name of an auxiliary indicator—namely “CHNG-CLI”, “CHNG-COVID”, “CTIS-CLI-in-community”, “DV-CLI”, or “Google-AA”—interchangeably with the model in forecasting, (3), or hotspot prediction, (5), that uses this same indicator as a feature (the meaning should be clear from the context). So, for example, the CHNG-CLI model in forecasting is the one in (3) that sets *X*_*ℓ,t*_ to be the value of the CHNG-CLI indicator at location *ℓ* and time *t*. Finally, we use the term “indicator model” to refer to any one of the ten models of the form (3) or (5) (five from each of the forecasting and hotspot prediction tasks).

Below is a summary of the high-level conclusions.

- Stratifying predictions by the ahead value (*a* = 7, …, 21), and aggregating results over the prediction period (early June through end of December 2020), we find that each of the indicator models generally gives a boost in predictive accuracy over the AR model, in both the forecasting and hotspot prediction tasks. The gains in accuracy generally attenuate as the ahead value grows.
- In the same aggregate view, CHNG-COVID and DV-CLI offer the biggest gains in both forecasting and hotspot prediction. CHNG-CLI is inconsistent: it provides a large gain in hotspot prediction, but little gain in forecasting (it seems to be hurt by a notable lack of robustness, due to backfill). CTIS-CLI-in-community and Google-AA each provide decent gains in forecasting and hotspot prediction. The former’s performance in forecasting is notable in that it clearly improves on AR even at the largest ahead values.
- In a more detailed analysis of forecasting performance, we find that the indicator models tend to be better than AR when case rates are flat or decreasing (most notable in CHNG-COVID and CTIS-CLI-in-community), but can be worse than AR when case rates are increasing (this is most notable in CHNG-CLI and DV-CLI). More rarely does an indicator model tend to beat AR when case rates are increasing, but there appears to be evidence of this for the Google-AA model.
- In this same analysis, when an indicator model performs better than AR in a decreasing period, this tends to co-occur with instances in which the indicator “leads” case rates (meaning, roughly, on a short-time scale in a given location, its behavior mimics that of future case rates). On the other hand, if an indicator model does better in periods of increase, or worse in periods of increase or decrease, its performance is not as related to leadingness.

Finally, to quantify the importance of training and making predictions using proper vintage data, we ran a parallel set of forecasting and hotspot prediction experiments using finalized data. The results, given in the Supplementary Materials, show that training and making predictions on finalized data can result in overly optimistic estimates of true test-time performance (up to 10% better in terms of average WIS or AUC). Furthermore, since indicators can have greatly different backfill profiles, the use of finalized data in retrospective evaluations changes the relative ranking of models. For example, CHNG-CLI and DV-CLI, when trained on finalized data, perform very similarly in forecasting. This makes sense since they are both claims-based indicators that are supposedly measuring the same thing. However, DV-CLI outperforms CHNG-CLI on vintage data, reflecting its has a less severe backfill profile.

The Supplement provides a number of other additional analyses; for example, we examine two assumption-lean methods for assessing the statistical significance of our main results.

Code to reproduce all results can be found at https://github.com/cmu-delphi/covidcast-pnas/tree/main/forecast/code.

### 2.1 Aggregate Results by Ahead Value

Figure 3 (left panel) displays evaluation results for forecasting, stratified by ahead value and averaged over all HRRs and forecast dates. Shown is the average WIS for each forecast model divided by that from a baseline model. The baseline model is a flat-line forecaster which forms its median forecast by using the most recent value *Y*_*ℓ,t*_ for all aheads *Y*_*ℓ,t*+*a*_, with predicted quantiles defined by the empirical distribution of the residuals from this median forecast over recent history. This is the same baseline model as in the COVID-19 Forecast Hub. Here, we use the baseline model in order to scale mean WIS to put it on an interpretable, unitless scale. In the figure, we can see that all curves are below 1, which means (smaller WIS is better) that all of the models, including AR, outperform the baseline on average over the forecasting period. On the other hand, the models deliver at best an improvement of about 20% in average WIS over the baseline model, with this gap narrowing to about 10% at the largest ahead values, illustrating the difficulty of the forecasting problem.

**Figure 3:**
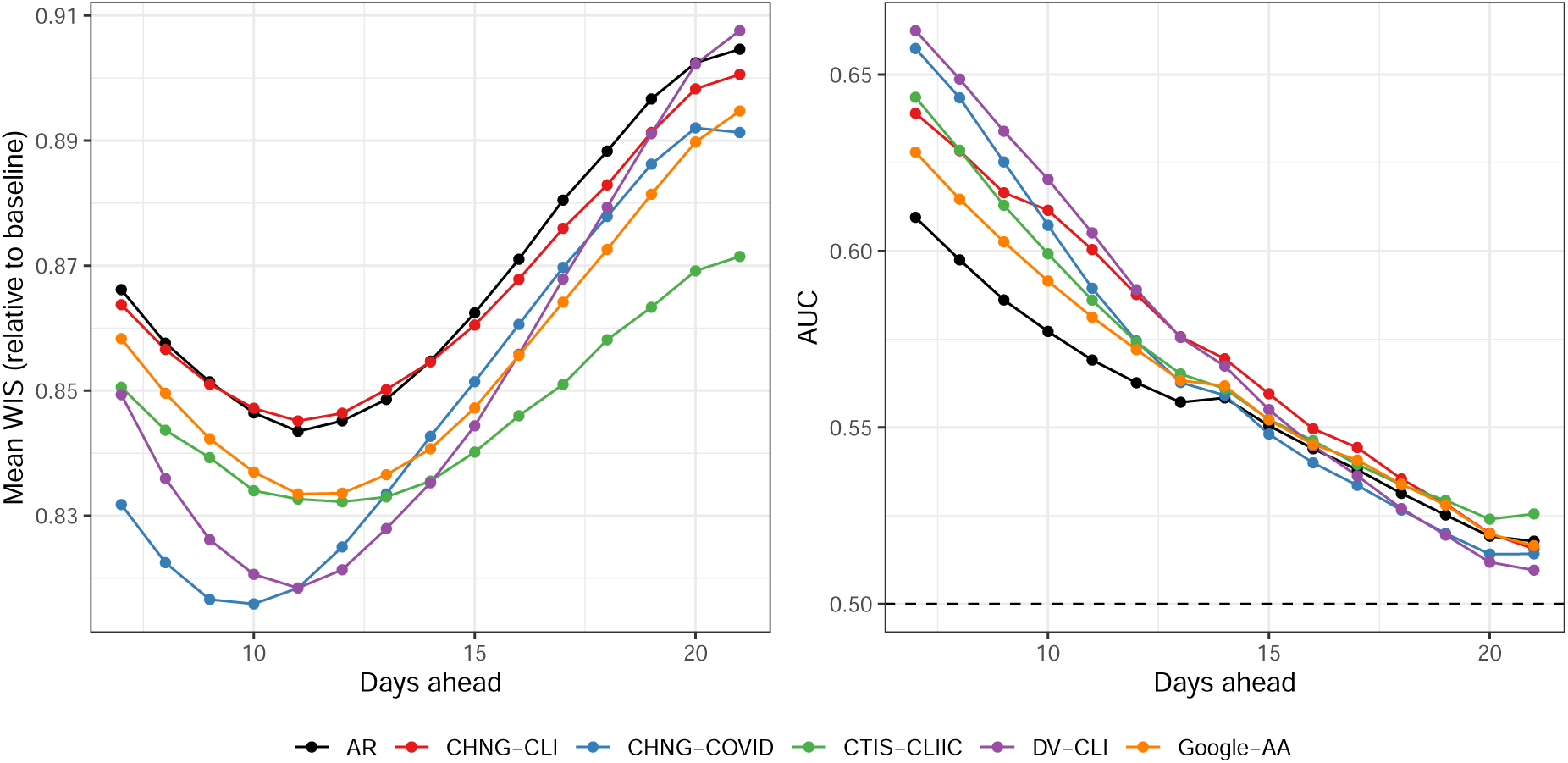
Main results for both tasks. Left: average WIS for each forecast model, over all forecast dates and all HRRs, divided by the average WIS achieved by a baseline model (a probabilistic version of the flat-line forecaster). Right: area under the curve for each hotspot prediction model, calculated over all prediction dates and all HRRs. Here and in all figures we abbreviate CTIS-CLI-in-community by CTIS-CLIIC.

We can also see from the figure that CHNG-COVID and DV-CLI offer the biggest gains over AR at small ahead values, followed by CTIS-CLI-in-community and Google-AA, with the former providing the biggest gains at large ahead values. The CHNG-CLI model performs basically the same as AR. This is likely due to the fact that CHNG-CLI suffers from volatility due to backfill. The evidence for this explanation is twofold: (1) the CHNG-CLI model benefits from a more robust method of aggregating WIS (geometric mean; shown in the Supplementary Materials); and (2) when we train and make predictions on finalized data, it handily beats AR, on par with the best-performing models (also shown in the Supplement).

Figure 3 (right panel) displays the results for hotspot prediction, again stratified by ahead value and averaged over all HRRs and prediction dates. We can see many similarities to the forecasting results (note: now larger AUC is better). For example, CHNG-COVID and DV-CLI offer the biggest improvement over AR, and all models, including AR, degrade in performance towards the baseline (in this context, a classifier based on random guessing, which achieves an AUC of 0.5) as the ahead values grow, illustrating the difficulty of the hotspot prediction problem. A clear difference, however, is that the CHNG-CLI model performs quite well in hotspot prediction, close to the best-performing indicator models for many of the ahead values. This may be because volatility in the CHNG-CLI indicator plays less of a role in the associated logistic model’s predicted probabilities (in general, a sigmoid function can absorb a lot of the variability in its input).

### 2.2 Implicit Regularization Hypothesis

One might ask if the benefits observed in forecasting and hotspot prediction have anything to do with the actual auxiliary indicators themselves. A plausible alternative explanation is that the indicators are just providing *implicit regularization* on top of the basic AR model, in the same way any noise variable might, if we were to use it to create lagged features in (3) and (5).

To test this hypothesis, we reran all of the prediction experiments but with *X*_*ℓ,t*_ in each indicator model replaced by suitable random noise (bootstrap samples from a signal’s history). The results, shown and explained more precisely in the Supplement, are vastly different (i.e., worse) than the original set of results. In both forecasting and hotspot prediction, the “fake” indicator models offer essentially no improvement over the pure AR model, which—informally speaking—strongly rejects the implicit regularization hypothesis.

On the topic of regularization, it is also worth noting that the use of *ℓ*_1_ regularization (tuned using cross-validation) in fitting any of the models in (1), (3), (4), or (5) did not generally improve their performance (experiments not shown). This is likely due to the fact that the number of training samples is large compared to the number of features (6,426 training samples and only 3–6 features).

### 2.3 Evaluation in Up, Down, and Flat Periods

The course of the pandemic has played out quite differently across space and time. Aggregating case rates nationally shows three pronounced waves, but the behavior is more nuanced at the HRR level. Figure 2 is a single example of a forecast in a period of relatively flat case trends, as New York City enters what would become its second wave. The AR forecaster’s 50% prediction interval contains this upswing, but its forecasted median is clearly below the finalized case data. Unfortunately, this behavior is fairly typical of all forecasters: during upswings, the forecasted median tends to fall below the target, while the reverse is true during downswings.

Figure 4 shows histograms of the differences in WIS of the AR model and each indicator model, where we stratify these differences by whether the target occurs during a period of increasing cases rates (up), decreasing case rates (down), or flat case rates (flat). To define the increasing period, we use the same definition we used for the hotspot task in Table 1. Therefore, all hotspots are labeled “up”, while all non-hotspots are either “flat” or “down”. For the “down” scenario, we simply use the opposite of the hotspot definition: *Y*_*ℓ,t*_ decreases by more than 20% relative to the preceding week.

**Figure 4:**
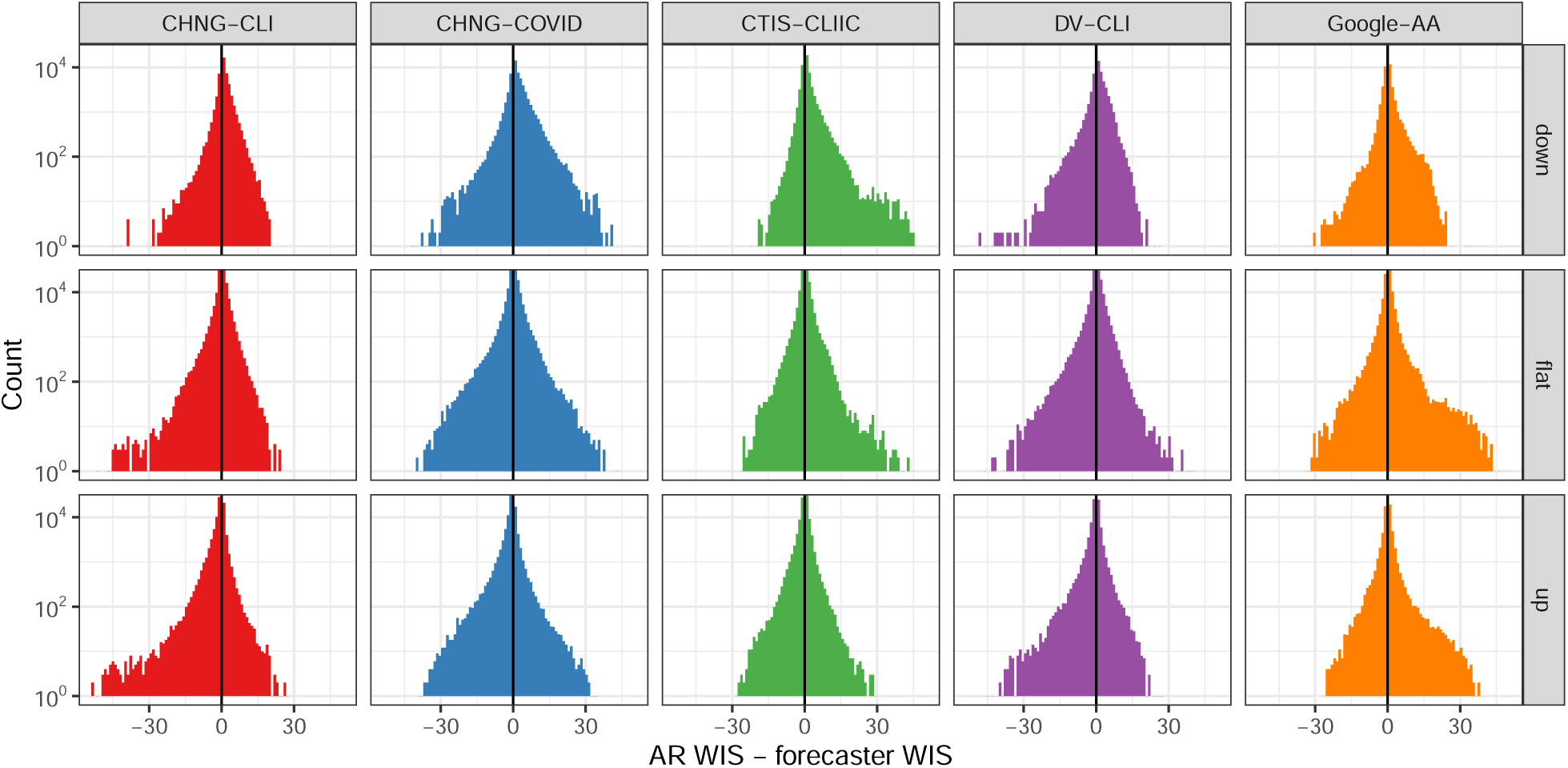
Histogram of the difference in WIS for the AR model and that for each indicator model, stratified by up, down, or flat period, measured in terms of case trends. Note that *larger* differences here are better for each indicator model. The y-axis is on the log scale to emphasize tail behavior.

While the performance of all models, including AR, generally degrades in up periods, different models exhibit different and interesting patterns. CHNG-CLI, CHNG-COVID, Google-AA, and especially CTIS-CLI-in-community have large right tails (showing improvements over AR) during the down periods. Google-AA and CTIS-CLI-in-community have large right tails during the flat periods. CHNG-CLI and DV-CLI have large left tails (poor forecasts relative to AR) in flat and up periods. Google-AA is the only model that outperforms the AR model, on average, in up periods. Overall, the indicators seem to help more during flat or down periods than during up periods, with the exception of Google-AA.

The Supplement pursues this analysis further. For example, we examine classification accuracy and log-likelihood for the hotspot task and find a similar phenomenon: the indicators considerably improve accuracy or log-likelihood during flat or down periods, with more mixed behavior during up periods when CHNG-CLI, CHNG-COVID, and DV-CLI, in particular, lead to decreased performance.

### 2.4 Effects of Leading or Lagging Behavior

As described in the methods section, each of the indicators we examine could be said to measure aspects of disease progression that would precede a positive test. That is, we imagine that these signals should “lead” cases. It is entirely reasonable to imagine that, prior to an increase of confirmed COVID-19 tests reported by a public health authority in a particular location, we would see an increase in medical insurance claims for COVID-related outpatient visits. However, it may well be the case that such behavior is different during different periods. In fact, we find empirically that the “leadingness” of an indicator (degree to which it leads case activity) tends to be more pronounced in down or flat periods than in up periods, a plausible explanation for the decreased performance in up periods noted above.

In the supplement, we define a quantitative score to measure the leadingness of an indicator, at any time *t* and any location *ℓ*, based on cross correlations to case rates over a short time window around *t*. The higher this score, the greater it “leads” case activity. This analysis is closely related to Granger causality [10] and draws on a large body of prior work that measures leadingness in economic time-series, e.g., [46–58]. Figure 5 displays correlations between the leadingness score of an indicator and the WIS difference (AR model minus an indicator model), stratified by whether the target is classified as up, down, or flat. One would naturally expect that the WIS difference would be positively correlated with leadingness. Somewhat surprisingly, this relationship turns out to be strongest in down periods and weakest in up periods. In fact, it is very nearly the case that for each indicator, the strength of correlations only decreases as we move from down to flat to up periods. In the supplement, we extend this analysis by studying analogous “laggingness” scores, but we do not find as clear patterns.

**Figure 5:**
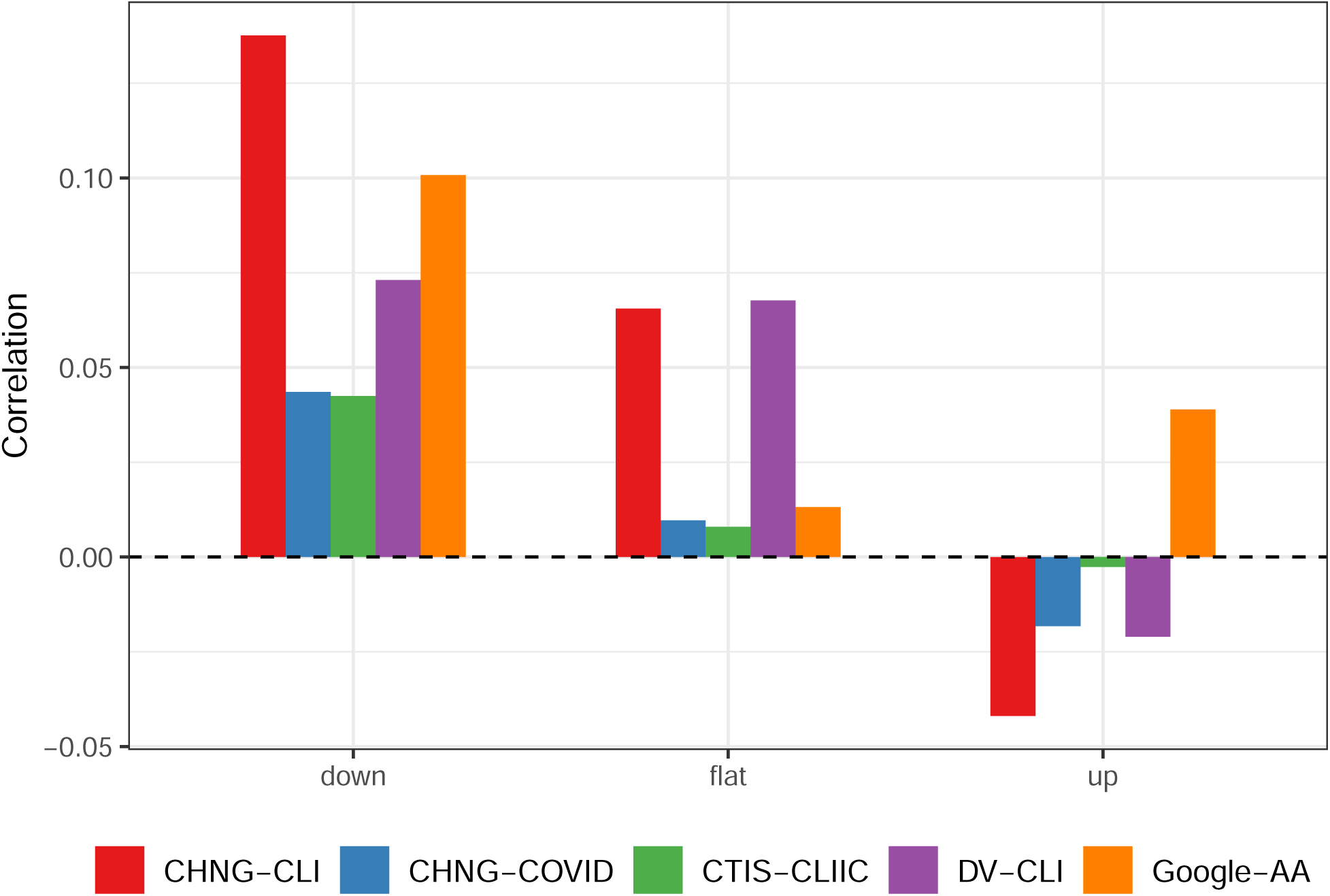
Correlation of the difference in WIS with the “leadingness” of the indicator at the target date, stratified by up, down, or flat period.

## 3 Discussion

Can auxiliary indicators improve COVID-19 forecasting and hotspot prediction models? Our answer, based on analyzing five auxiliary indicators from the COVIDcast API (defined using from medical insurance claims, internet-based surveys, and internet search trends) is undoubtedly “yes”. However, there are levels of nuance to such an answer that must be explained. None of the indicators that we have investigated are transformative, rendering the prediction problem easy when it was once hard (in the absence of auxiliary information). Rather, the gains in accuracy from the indicator models (over an autoregressive model based only on past case rates) appear to be nontrivial, and consistent across modes of analysis, but modest. In forecasting, the indicator models are found to be most useful in periods in which case rates are flat or trending down, rather than periods in which case rates are trending up (as one might hope to see is the benefit being provided by a hypothetical “leading indicator”).

As described previously, it is likely that we could improve the indicator models by using location-specific effects, as well as using nowcasting techniques to estimate finalized indicator values before we use them as features (to account for backfill in the claims-based signals in particular). Beyond this, it is certainly possible that more sophisticated models for forecasting or hotspot prediction would lead to different results, and possibly even different insights. Natural directions to explore include using multiple indicators in a single model, allowing for interaction terms, and leveraging HRR demographics or mobility patterns. That said, we are doubtful that more sophisticated modeling techniques would change the “topline” conclusion—that auxiliary indicators can provide clear but modest gains in forecasting and hotspot prediction.

However, rigorously vetting the details for more sophisticated models, as well as the generalizability of our findings to different geographic resolutions, both remain important directions for future study. For example, in the Supplementary Materials we show that for forecasting at the state level, the benefits of including indicators in the AR model are generally less clear (compared to those observed at the HRR level). A plausible explanation: at the state level, where the signal-to-noise ratio (SNR) is higher, AR performs better overall and represents a higher standard (when asking whether it can be improved upon using the indicators). At the HRR level, where the SNR is lower, including the indicators as additional linear features in the AR model probably delivers a kind of variance reduction (just like averaging independent terms would) which contributes to improved accuracy. But as the SNR increases, this variance reduction becomes less important, and perhaps we must use more sophisticated modeling techniques to extract comparable value from the indicators.

We reiterate the importance of using vintage data for rigorous backtesting. Data sources that are relevant to public health surveillance are often subject to revision, sometimes regularly (as in medical claims data) and sometimes unpredictably (such as COVID-19 case reports). When analyzing models that are designed to predict future events, if we train these models and make predictions using finalized data, then we are missing a big part of the story. Not only will our sense of accuracy be unrealistic, but certain models may degrade by a greater or lesser extent when they are forced to reckon with vintage data, so backtesting on finalized data may lead us to make modeling decisions that are suboptimal for true test-time performance.

In this paper, we have chosen to consider only very simple forecasting models, while devoting most of our effort to accounting for as much of the complexity of the underlying data and evaluation as possible. In fact, our paper is not only about providing rigorous answers to questions about model comparisons in COVID-19 forecasting and hotspot prediction, but also about demonstrating how one might go about answering such questions in general. We hope that others will leverage our work, and build on it, to guide advances on the frontier of predictive modeling for epidemics and pandemics.

## Data Availability

All data is publicly available from the linked URL to the COVIDcast Epidata API.

https://cmu-delphi.github.io/delphi-epidata/api/covidcast.html

## Acknowledgements

We thank Matthew Biggerstaff, Logan Brooks, Johannes Bracher, Michael Johansson, Evan Ray, Nicholas Reich, and Roni Rosenfeld for several enlightening conversations about forecasting, scoring, and evaluation. This material is based on work supported by gifts from Facebook, Google.org, the McCune Foundation, and Optum; Centers for Disease Control and Prevention (CDC) grant U01IP001121; the Canadian Statistical Sciences Institute; National Sciences and Engineering Research Council of Canada (NSERC) grant RGPIN-2021-02618; and National Science Foundation Graduate Research Fellowship Program (NSF GRFP) award DGE1745016.

## Supplemental information

### A Finalized Versus Vintage Data

The goal of this section is to quantify the effect of not properly accounting for the question of “what was known when” in performing retrospective evaluations of forecasters. Figures 6 and 7 show what Figures 3 and 4 in the main paper would have looked like if we had simply trained all models using the finalized data rather than using vintage data. This comparison can be seen more straightforwardly in Figures 8 and 9, which show the ratio in performance between the vintage and finalized versions. When methods are given the finalized version of the data rather than the version available at the time that the forecast would have been made, all methods appear (misleadingly) to have better performance than they would have had if run prospectively. For example, for forecasting case rates 7-days ahead, the WIS of all methods is at least 8% larger than what would have been achieved using finalized data. This effect diminishes as the forecasting horizon increases, reflecting the fact that longer-horizon forecasters rely less heavily on recent data than very short-horizon forecasters. Crucially, some methods are “helped” more than others by the less scrupulous retrospective evaluation, underscoring the difficulty of avoiding misleading conclusions when performing retrospective evaluations of forecasters.

**Figure 6:**
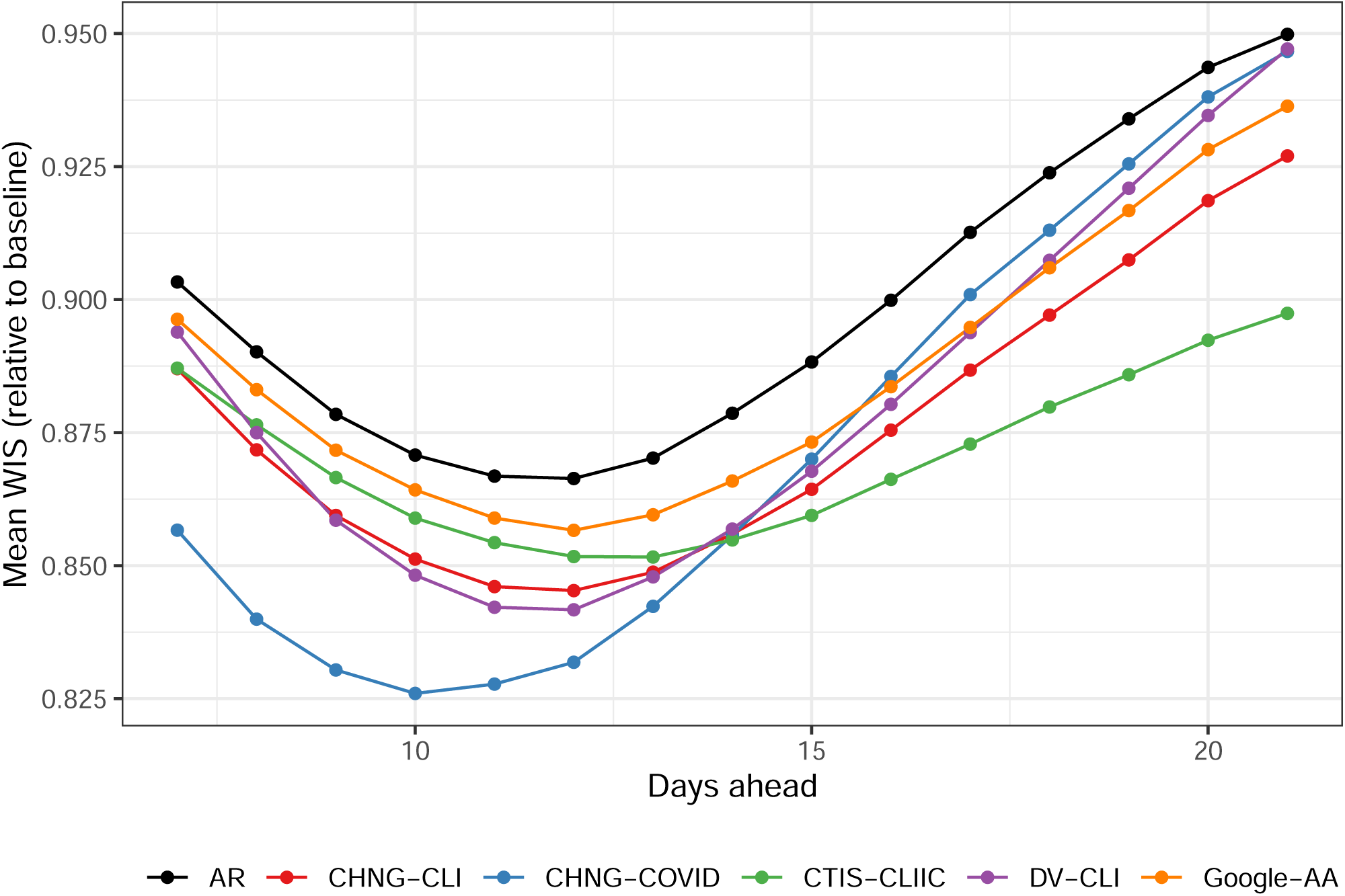
Forecasting performance using finalized data. Compare to Figure 3 in the manuscript.

**Figure 7:**
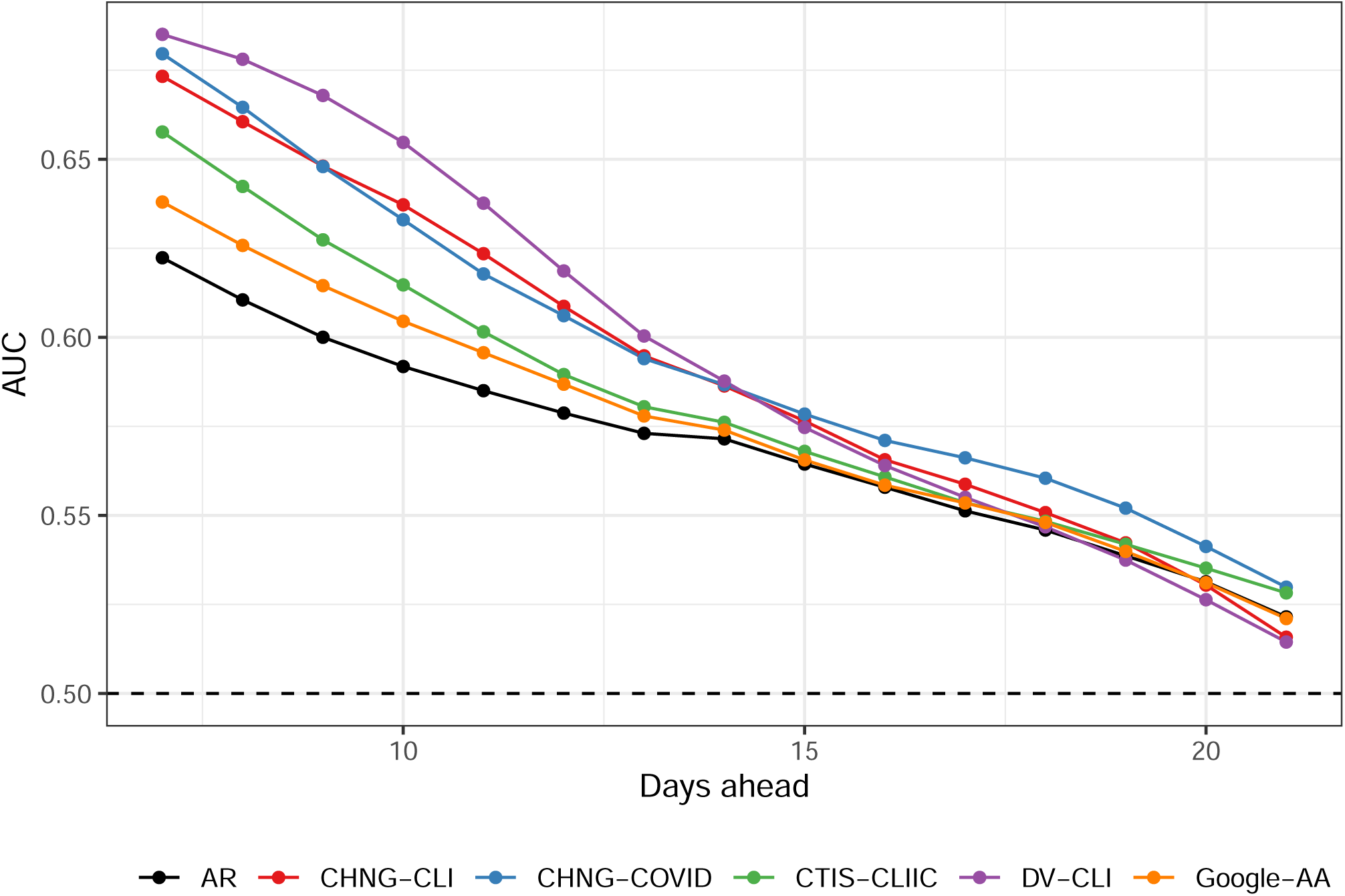
Hotspot prediction performance using finalized data. Compare to Figure 4 in the manuscript.

**Figure 8:**
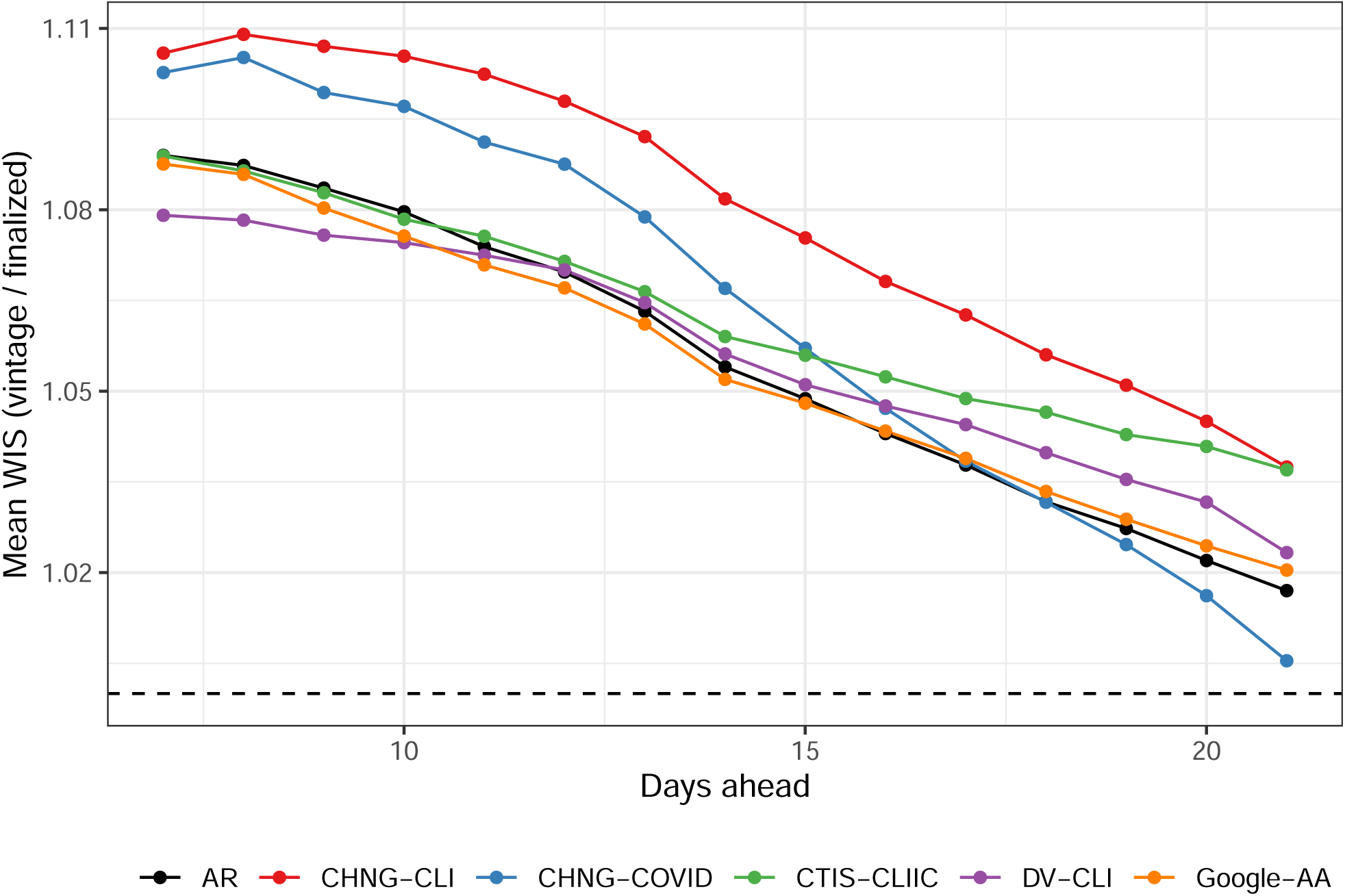
Forecast performance with vintage compared to finalized data. Using finalized data leads to overly optimistic performance.

**Figure 9:**
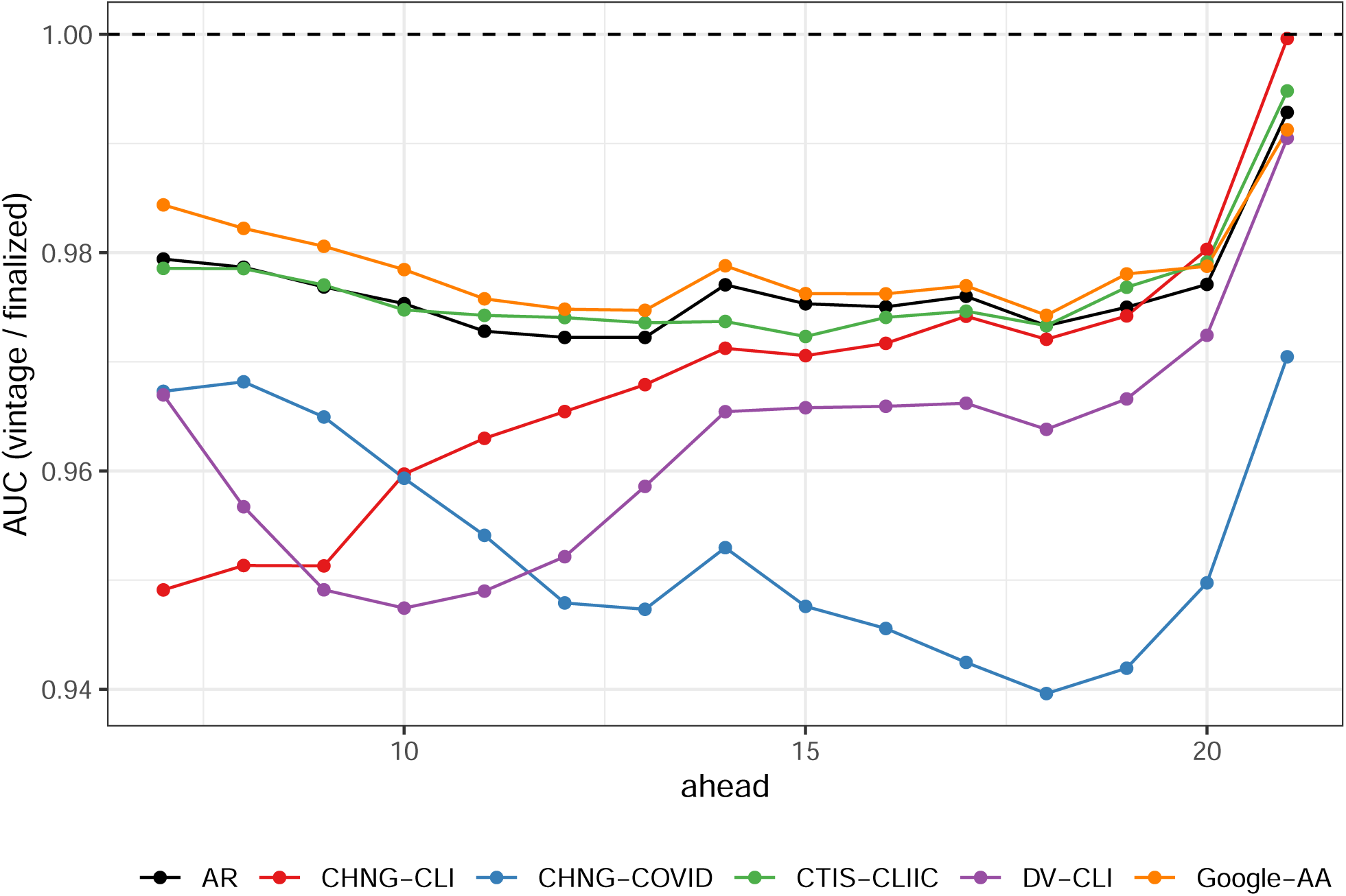
Hotspot prediction performance with vintage compared to finalized data. Using finalized data leads to overly optimistic performance.

CHNG-CLI (and, to a lesser extent, the other claims-based signals) is the most affected by this distinction, reflecting the latency in claims-based reporting. This highlights the importance of efforts to provide “nowcasts” for claims signals (which corresponds to a 0-ahead forecast of what the claims signal’s value will be once all data has been collected). Looking at the CHNG-CLI and DV-CLI curves in Figure 6, we can see that they perform very similarly when trained on the finalized data. This is reassuring because they are, in principle, measuring the same thing (namely, the percentage of outpatient visits that are primarily about COVID-related symptoms), but based on data from different providers. The substantial difference in their curves in Figure 3 of the main paper must, therefore, reflect their having very different backfill profiles.

While using finalized rather than vintage data affects DV-CLI the least for forecasting, it is one of the most affected methods for the hotspot problem. This is a reminder that the forecasting and hotspot problems are fundamentally different problems. For example, the hotspot problem does not measure the ability to distinguish between flat and downward trends.

Even the AR model is affected by this distinction, reflecting the fact that the case rates themselves (i.e., the response values) are also subject to revision. The forecasters based on indicators are thus affected both by revisions to the indicators and by revisions to the case rates. And in the case of the Google-AA model, in which we only used finalized values for the Google-AA indicator, the difference in performance can be wholly attributed to revisions of case rates.

### B Robust Aggregation

In this section, we consider using the geometric mean instead of the usual (arithmetic) mean when aggregating the weighted interval score (WIS) across location-time pairs. Aside from the geometric mean being generally more robust to large values, there are two reasons why using it may be desirable.

1. WIS is right-skewed, being bounded below by zero and having occasional very large values. Figure 10 illustrates that the densities appear roughly log-Gaussian. The geometric mean is a natural choice in such a context since it can be viewed as a measure of centrality on the log scale (it is the exponential of the arithmetic mean of log values).
2. In the main paper, we report the ratio of the mean WIS of a forecaster to the mean WIS of the baseline forecaster. Another choice could be to take the mean of the ratio of WIS values for the two methods. This latter choice would penalize a method less for doing poorly where the baseline forecaster also does poorly.^1^ Using instead the geometric mean makes the order of aggregation and scaling immaterial since the ratio of geometric means is the same as the geometric mean of ratios.

**Figure 10:**
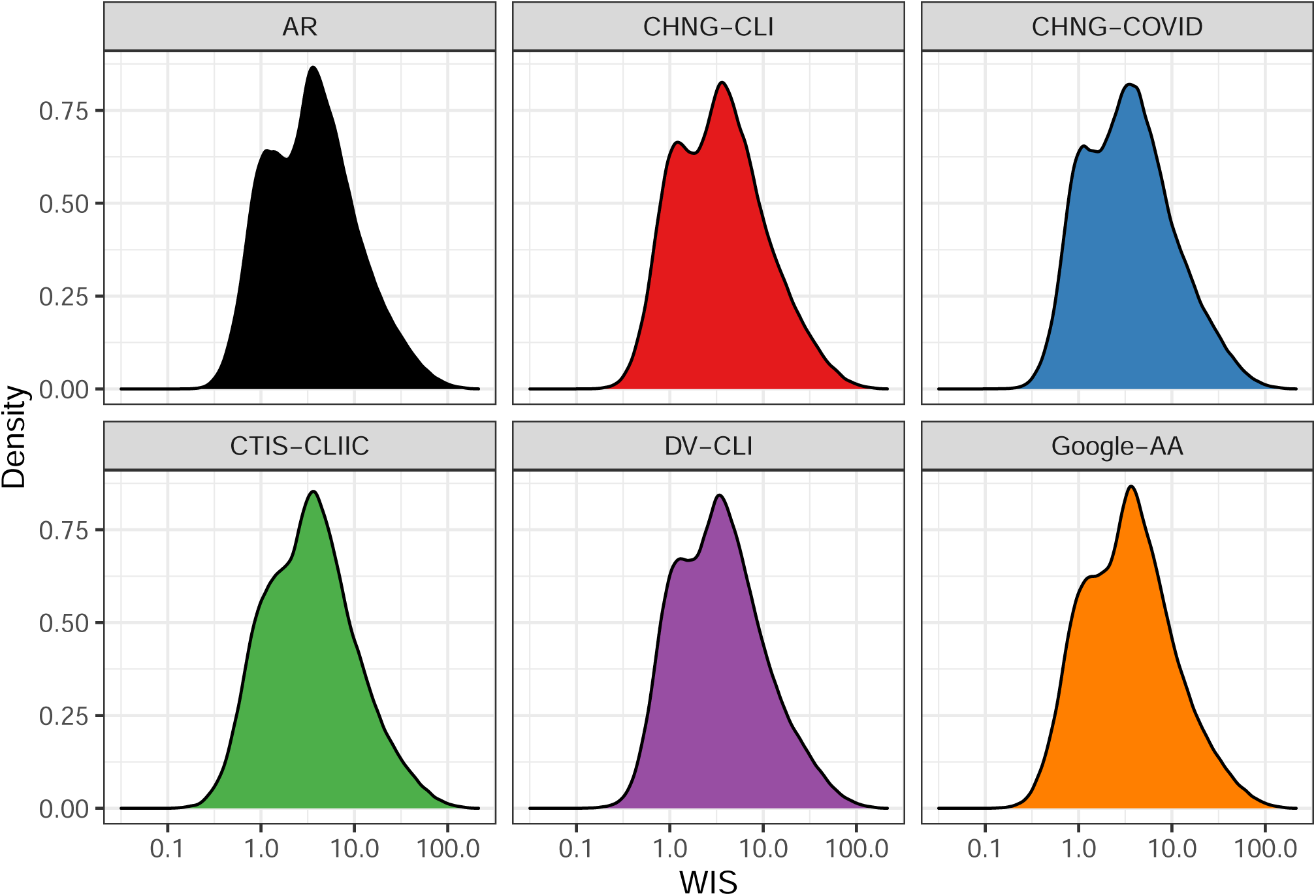
WIS values from forecast models, which appear to be roughly log-Gaussian.

Figure 11 uses the geometric mean for aggregation. Comparing this with Figure 3 in the main paper, we see that the main conclusions are largely unchanged; however, CHNG-CLI now appears better than AR. This behavior would be expected if CHNG-CLI’s poor performance is attributable to a relatively small number of large errors (as opposed to a large number of moderate errors). Indeed, Figure 5 of the main paper further corroborates this, in which we see the heaviest left tails occur for CHNG-CLI.

**Figure 11:**
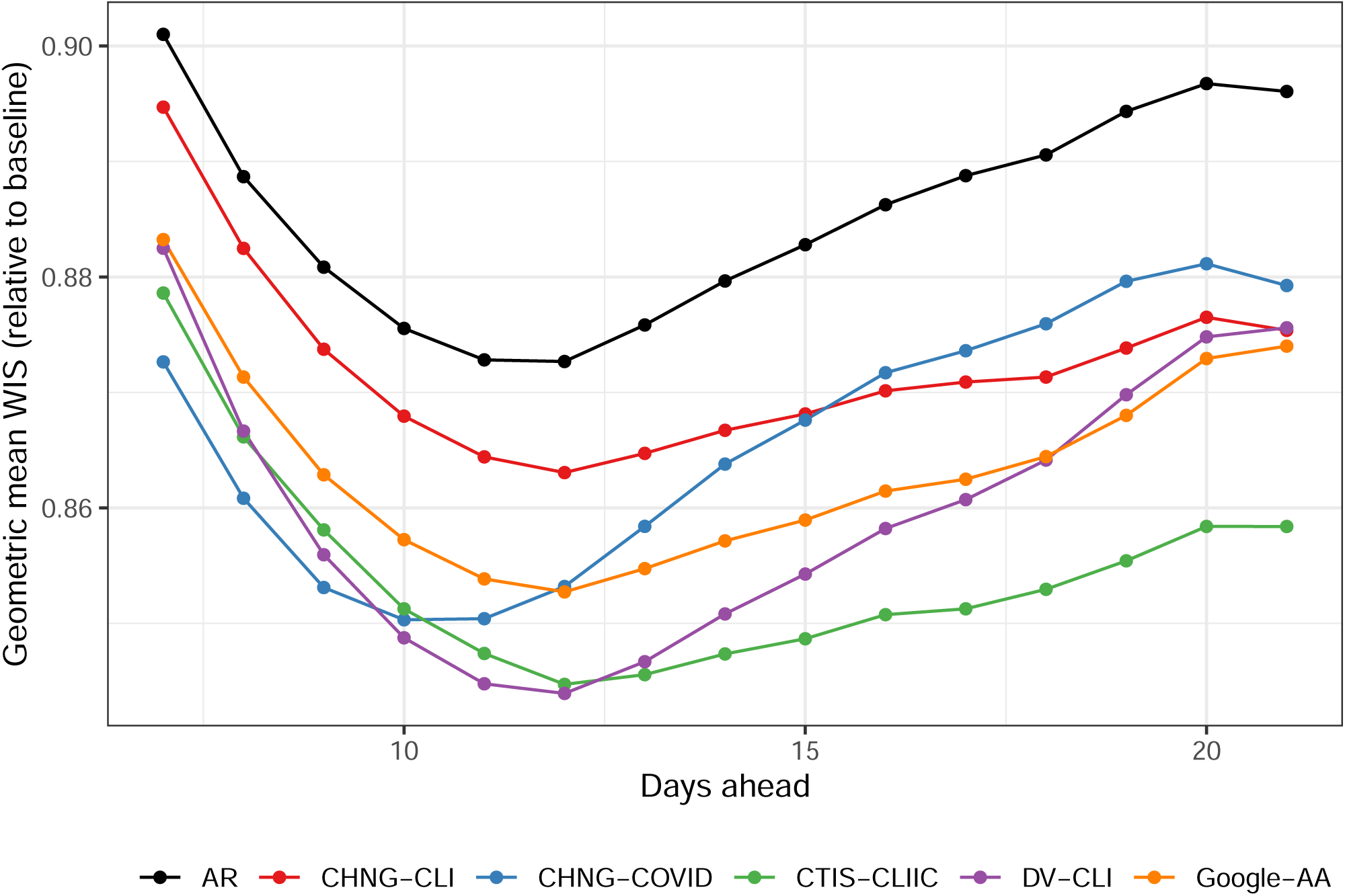
Forecast performance (using vintage data), summarized by geometric mean.

### C Comparing COVID-19 Forecast Hub Models

Since July of 2020, modelers have been submitting real-time forecasts of COVID-19 case incidence to the COVID-19 Forecast Hub [7]. This (along with forecasts of hospitalizations and deaths collected in the same Hub) serves as the source of the CDC’s official communications on COVID forecasting.

Our goal in this section is to compare the AR model and indicator models to those in the Hub, in terms of forecast errors aggregated over the same forecast tasks, to give a clear sense for how robust and effective the models we choose to investigate in the paper are relative to those in common operational use. This was prompted by a question from an anonymous reviewer of this paper, who asked why we chose to build our analysis of indicator utility around the AR model in the first place, and why we did not build it around others (say, the SIR model or more complex mechanistic models of disease transmission) that have occupied more of the spotlight over the course of the pandemic. The analysis presented here corroborates the claim that, while simple, the AR model, properly trained—using a quantile loss to directly estimate multiple conditional quantiles, a trailing training window of 21 days, pooling across all locations jointly, and fitting to case rates rather than counts (as we do in all our models in the main paper)—can be robust and effective, performing competitively to the top models submitted to the COVID-19 Forecast Hub, including the Hub’s ensemble model.

The closest forecast target in the Hub to that used in the main paper is state-level case incidence over an epiweek—defined by the sum of new case counts reported between a Sunday and the following Saturday (inclusive). Our forecast target, recall, is a 7-day trailing average of COVID-19 case incidence rates at the HRR level, which is different in three regards:

1. temporal resolution (daily versus weekly);
2. geographic resolution (HRRs versus states);
3. scale (rates versus counts).

While the first and third of these differences could be easily addressed post hoc—meaning, we can take always take our model’s output and multiply it by 7 in order to track the incidence over any given trailing week, and rescale it per location by population to bring it to the count scale—the second difference is not easy to adjust post hoc due to nonlinearity of the quantiles (a quantile of a linear combination of random variables is not simply the linear combination of their quantiles, but rather, depends intricately on the correlations between the random variables).

Therefore, to make the comparison to models in the Hub as direct as possible, we retrained our models over the same forecast period as in the main paper, and with the same general setup entirely, except at the state rather than HRR level. We then rescaled them post hoc to account for the different temporal resolution and the rate-versus-count scaling (first and third points in the above list). The results are given in Figure 12. The evaluation was carried out exactly as in the main paper, and the figure displays both mean WIS and geometric mean WIS, as a function of ahead, relative to the baseline model. Furthermore, to account for missingness (not all teams submitted forecasts to the Hub for all locations and ahead values for the entire period), we first dropped any forecaster than submitted for less than 6 weeks, and then restricted the aggregation metrics (mean or geometric mean) to errors from commonly available forecast tasks.

**Figure 12:**
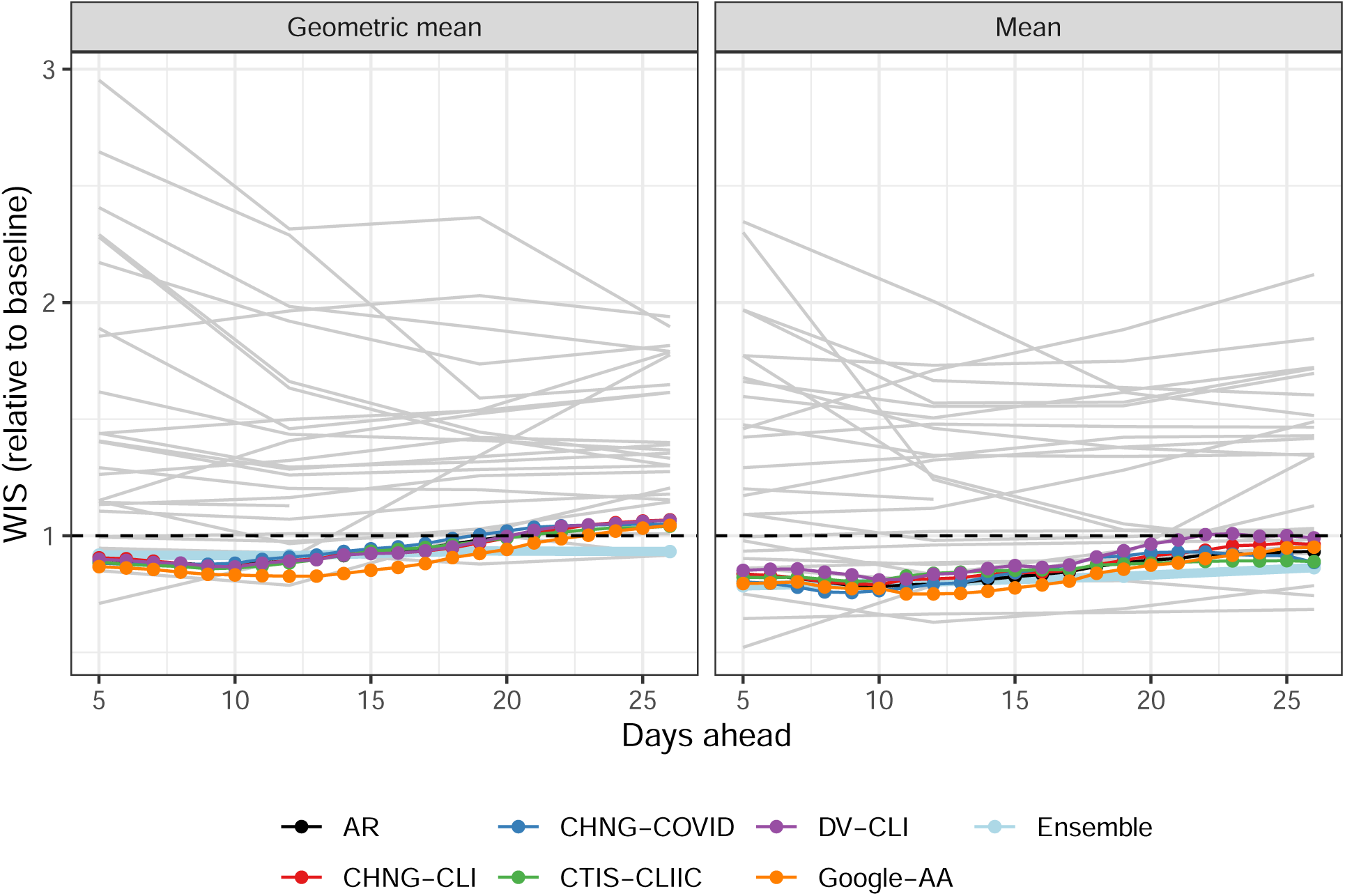
Forecast performance for AR and indicator models, each retrained at the state level, compared to models submitted to the COVID-19 Forecast Hub over the same period. The thin grey lines are individual models from the Hub; the light blue line is the Hub ensemble model. To align prediction dates as best as possible, we look at the AR and indicator model forecasts for 5, 12, 19, and 26 days ahead; this roughly corresponds to 1, 2, 3, and 4 weeks ahead, respectively, since in the Hub, models typically submit forecasts on a Tuesday for the epiweeks aligned to end on each of the following 4 Saturdays.

By either metric, mean or geometric mean WIS relative to baseline, we can see in Figure 12 that the AR model examined in this paper is competitive with top models in the Hub, even outperforming the Hub ensemble model for smaller ahead values. The same general conclusion can be drawn for the indicator-assisted models as well. However, interestingly, a close inspection reveals that the AR model here is for the most part in the “middle of the pack” when compared to the indicator models, and only the Google-AA model offers clear improvement over AR for all aheads. This is likely due to the fact that at the state level, the signal-to-noise ratio (SNR) is generally higher, and AR model provides a higher standard (on which to expect improvement using an auxiliary indicator), since it is able to extract a clearer signal from past lags of case rates. At the HRR level, with lower SNR, using indicators as simple additional linear features in the AR model probably leads to a variance reduction that is enough to boost accuracy, but at the state level, perhaps more sophisticated modeling techniques are needed to extract comparable value from some of the indicators.

### D Statistical Significance

In the introduction of the main manuscript, we gave some reasons that we avoid making formal statements about statistical significance, preferring instead to examine the stability of our results in different contexts. There are strong reasons to avoid model-based significance tests because the necessary assumptions about stationarity, independence, and the model being true (or at least approximately true) are certainly violated. With those caveats in mind, we undertake two relatively assumption-lean investigations in this section. The first is a sign test for whether the difference between the AR model’s relative WIS and each other model’s relative WIS is centered at zero. (Relative WIS here means scaled by the WIS of the baseline model.) To mitigate the dependence across time (which intuitively seems to matter more than that across space), we computed these tests in a stratified way, where for each forecast date we run a sign test on the scaled errors between two models over all 306 HRRs. The results are plotted as histograms in Figure 13. For this figure, we use the total relative WIS over all aheads, but the histograms are largely similar for individual target horizons. If there were no difference, we would expect to see a uniform distribution. However, for each indicator model, we see many more small p-values than would be expected if the null hypothesis (that the AR model is better) were true.

**Figure 13:**
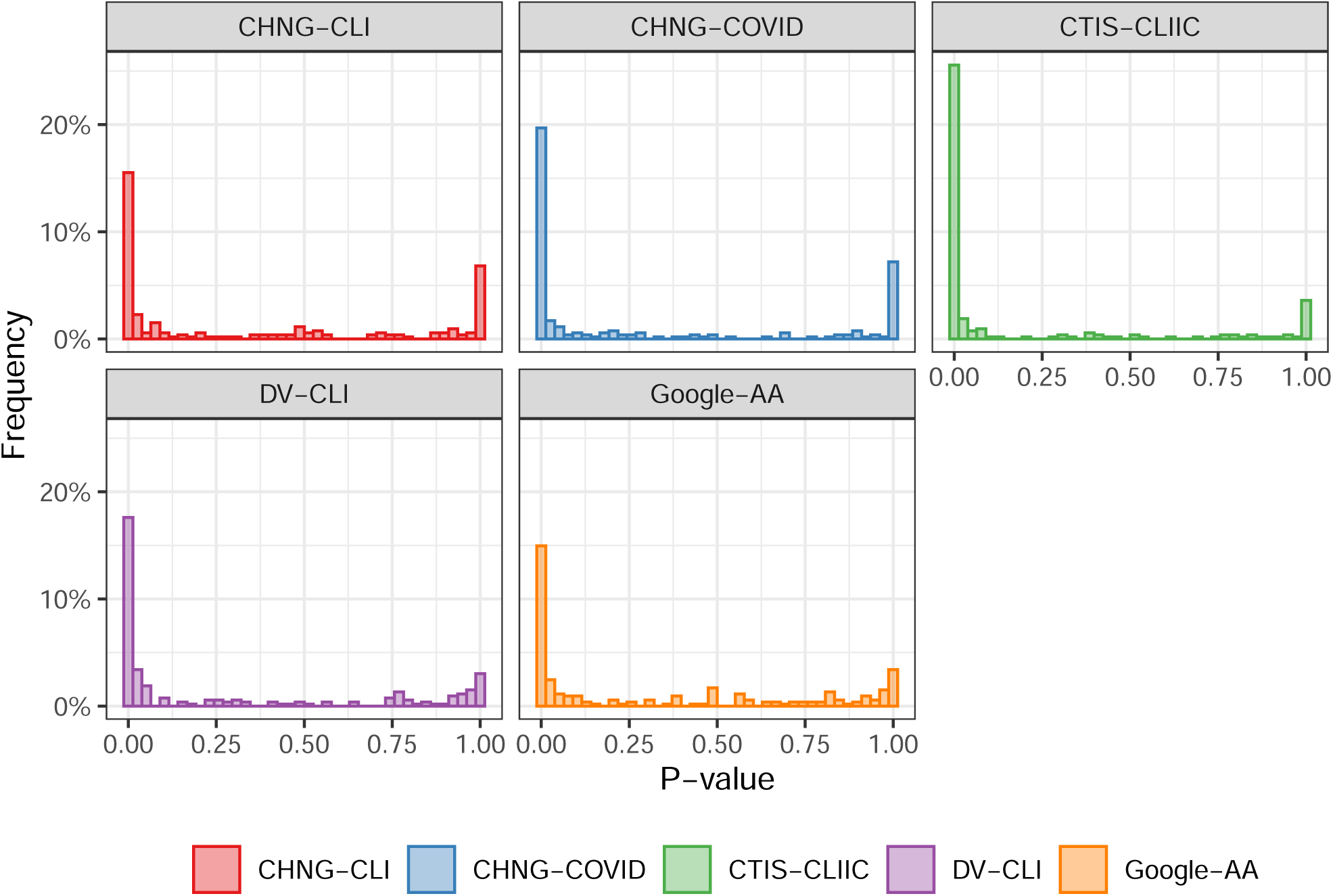
P-values from a one-sided sign test for improved forecast performance of the indicator-assisted models. Each p-value corresponds to a forecast date. The alternative hypothesis is that the AR model is better (median difference between the relative WIS of the AR model and an indicator model is negative).

Another relatively assumption-lean method of testing for differences in forecast accuracy is the Diebold-Mariano (DM) test [23, 25, 59]. Essentially, the differences between forecast errors are assumed to have a constant mean and a covariance that depends on time. Under these conditions, the asymptotic distribution for the standardized mean of the differences is limiting normal provided that a heteroskedasticity and autocorrelation robust estimate of the variance is used. Using the error as WIS across all HRRs and horizons (7 to 21 days ahead), we perform the DM test using both the mean relative to the baseline (as reported in the manuscript) and the geometric mean relative to the baseline as described above. The first two rows of Table 2 displays p-values for the test that each indicator model is no better than the AR model. In only a few instances—geometric mean and mean for the CHNG-CLI model, and mean for the CHNG-COVID model—do the p-values exceed conventional statistical significance thresholds.

**Table 2:** P-values from a one-sided Diebold-Mariano test for improvemed forecast performance when adding the indicators. The alternative hypothesis is that the AR model is better.

| metric | CHNG-CLI | CHNG-COVID | CTIS-CLIIC | DV-CLI | Google-AA |
| --- | --- | --- | --- | --- | --- |
| Geometric mean | 0.072 | 0.036 | 0.005 | 0.032 | 0.026 |
| Mean | 0.177 | 0.132 | 0.006 | 0.092 | 0.103 |

### E Bootstrap Results

As explained in Section 2.B of the main paper, a (somewhat cynical) hypothesis for why we see benefits in forecasting and hotspot prediction is that the indicators are not actually providing useful information but they are instead acting as a sort of “implicit regularization,” leading to shrinkage on the autoregressive coefficients and therefore to less volatile predictions. To investigate this hypothesis, we consider fitting “noise features” that in truth have zero relationship to the response. Recall (from the main paper) that at each forecast date, we train a model on 6,426 location-time pairs. Each indicator model uses 6 features, corresponding to the 3 autoregressive terms and the 3 lagged indicator values. To form noise indicator features, we replace their values with those from a randomly chosen time-space pair (while keeping the autoregressive features fixed). In particular, at each location *ℓ* and time *t*, for the forecasting task we replace the triplet (*X*_*ℓ,t*_, *X*_*ℓ,t*−7_, *X*_*ℓ,t*−14_) in Eq. (3) of the main paper with the triplet 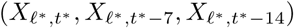, where (*ℓ*^*\**^, *t*^*\**^) is a location-time pair sampled with replacement from the 6,426 total location-time pairs. Likewise in the hotspot prediction task, we replace the triplet 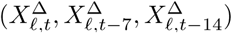 in Eq. (5) of the main paper with 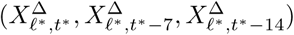 Figures 14–16 show the results. No method exhibits a noticeable performance gain over the AR method, leading us to dismiss the implicit regularization hypothesis.

**Figure 14:**
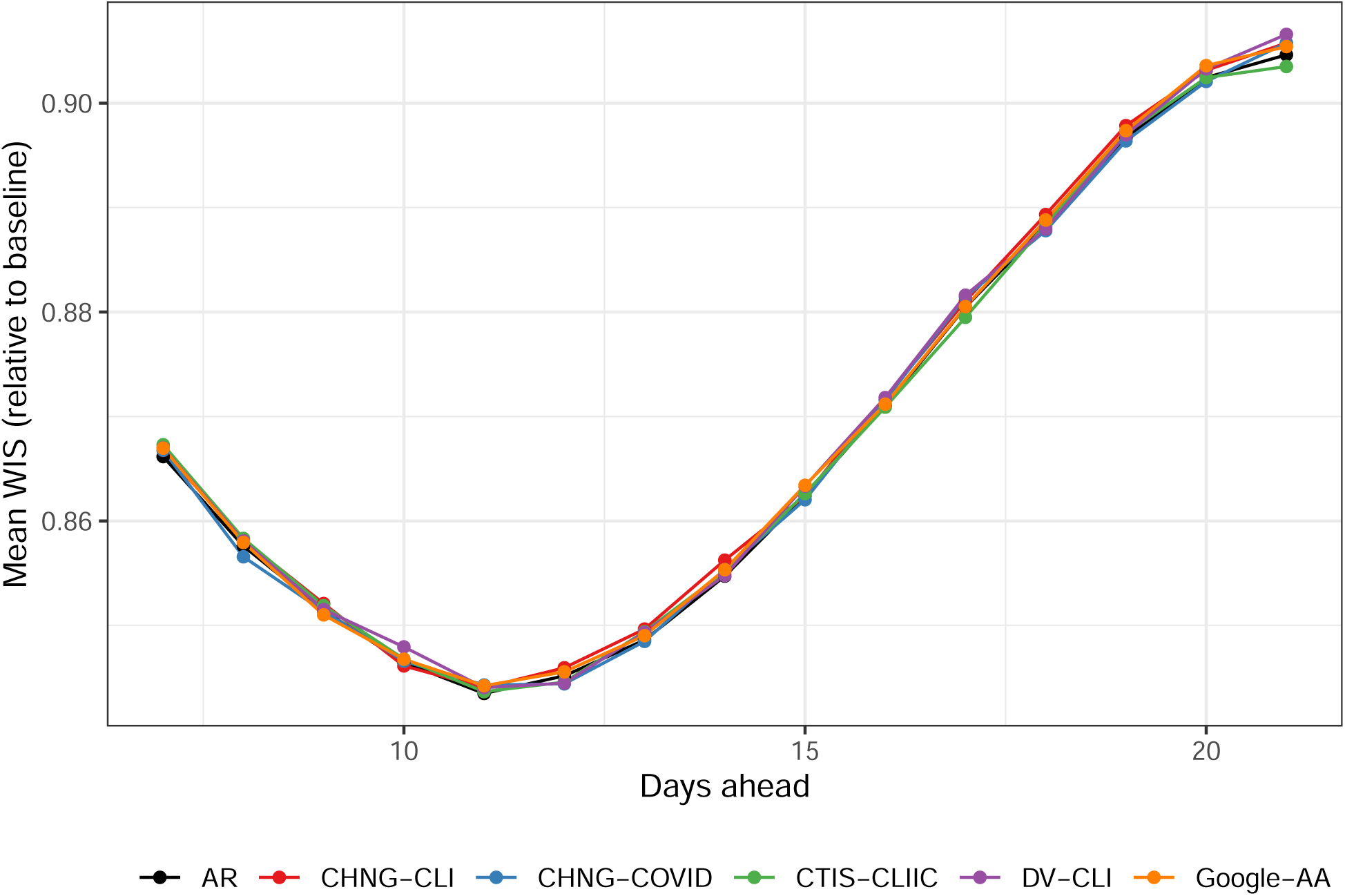
Forecast performance when indicators are replaced with samples from their empirical distribution. Performance is largely similar to the AR model.

**Figure 15:**
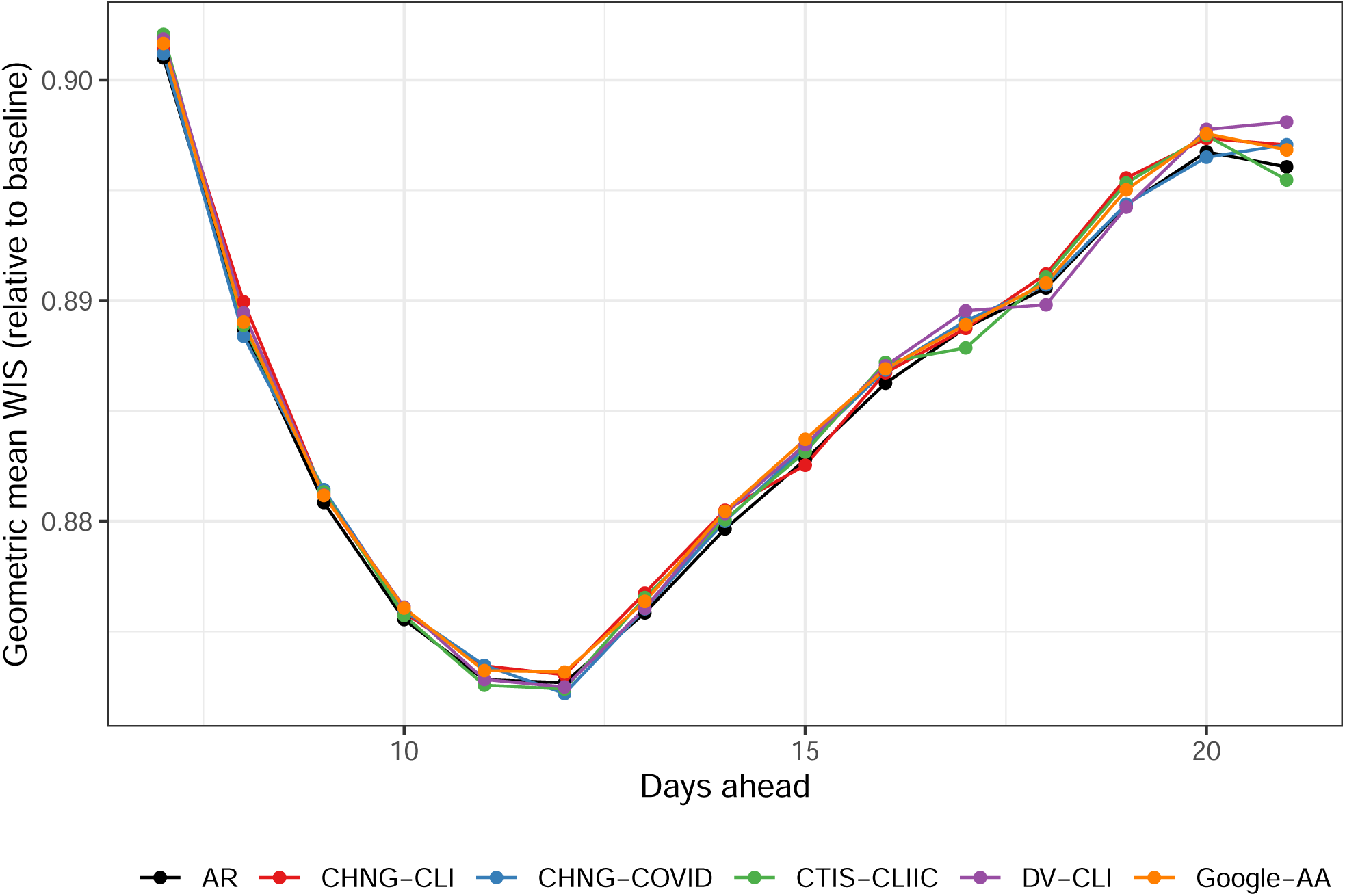
Forecast performance as measured with the geometric mean when indicators are replaced with samples from their empirical distribution. Performance is largely similar to the AR model.

**Figure 16:**
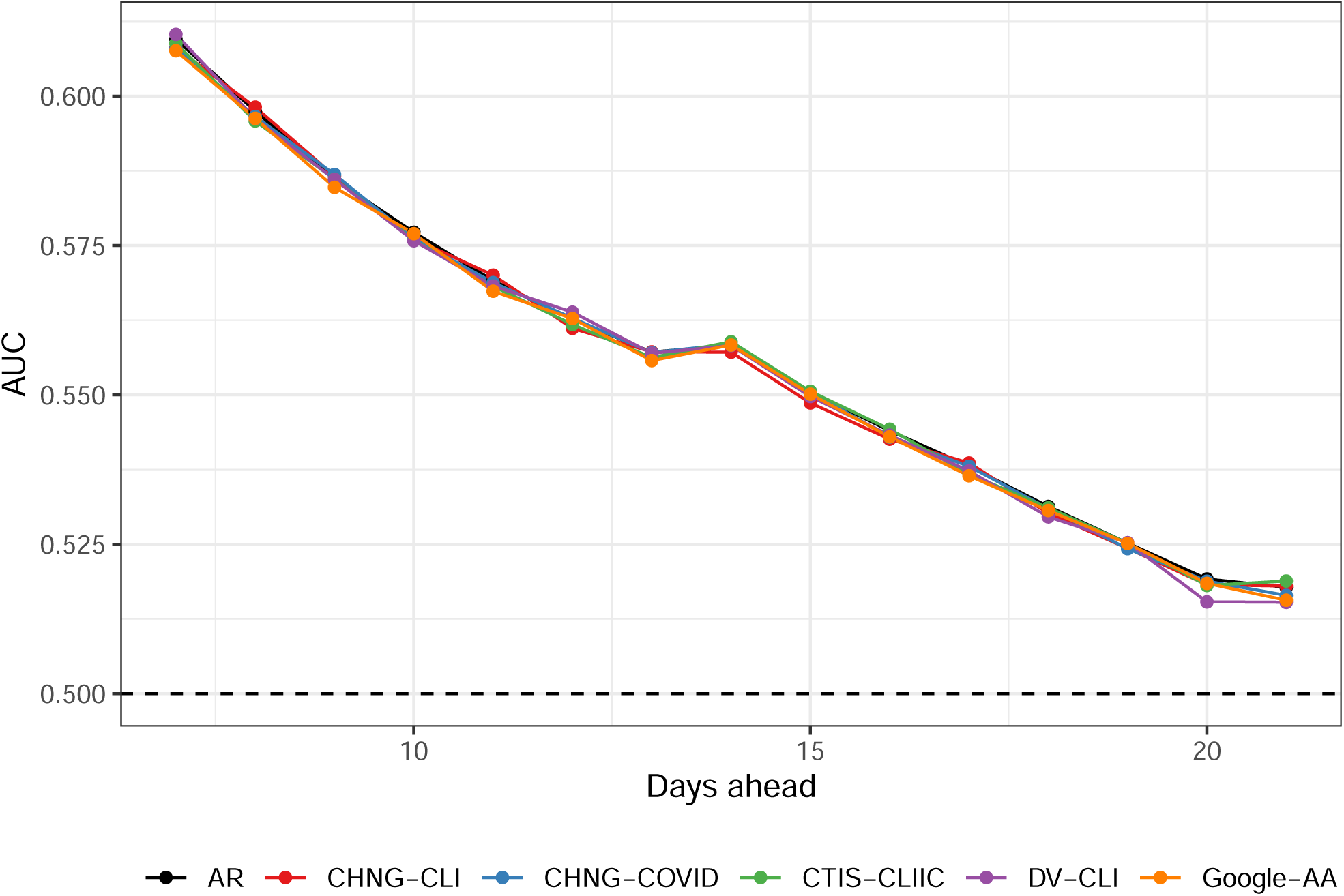
Hotspot prediction performance when indicators are replaced with samples from their empirical distribution. Performance is largely similar to the AR model.

### F Upswings and Downswings

In this section, we provide extra details about the upswing/downswing analysis described in the main manuscript, Section 2.C. Figure 17 shows the overall results, examining the average difference WIS(AR) − WIS(*F*) for each forecaster *F*, in each in each period. Figure 18 shows the same information for the hotspot task. On average, during downswings and flat periods, the indicator models have lower classification error and higher log likelihood than the AR model. For hotspots, both Google-AA and CTIS-CLIIC perform better than the AR model during upswings, in contrast to the forecasting task, where only Google-AA improves. In a related analysis, Figure 19 shows histograms of the Spearman correlation (Spearman’s *ρ*, a rank-based measure of association) between the ratio WIS(*F*)*/*WIS(AR) and the magnitude of the swing. Again we see that case rate increases are positively related to diminished performance of the indicator models.

**Figure 17:**
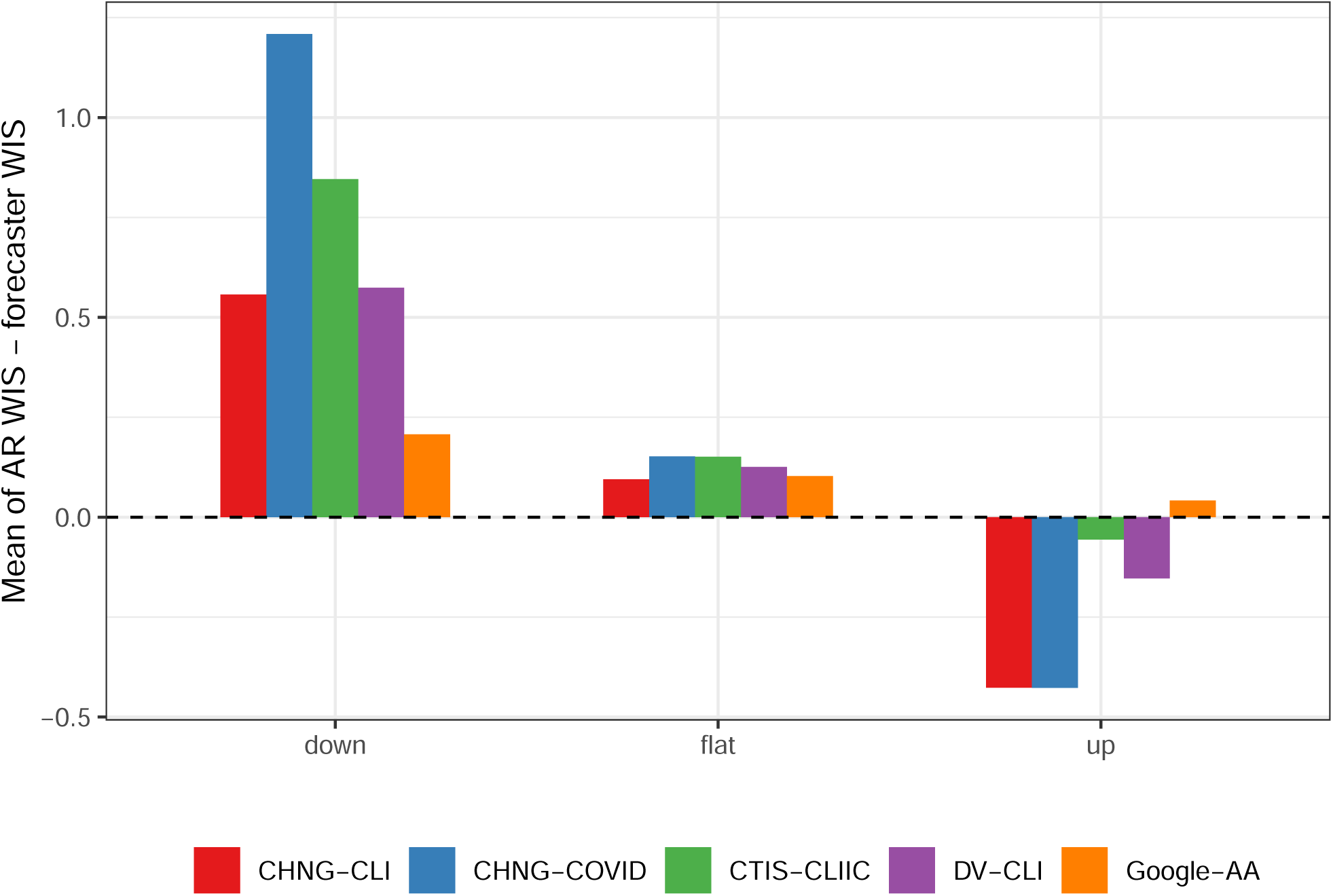
Average difference between the WIS of the AR model and of the indicator models, separated into up, down, and flat periods. The indicator models generally do best during down and flat periods.

**Figure 18:**
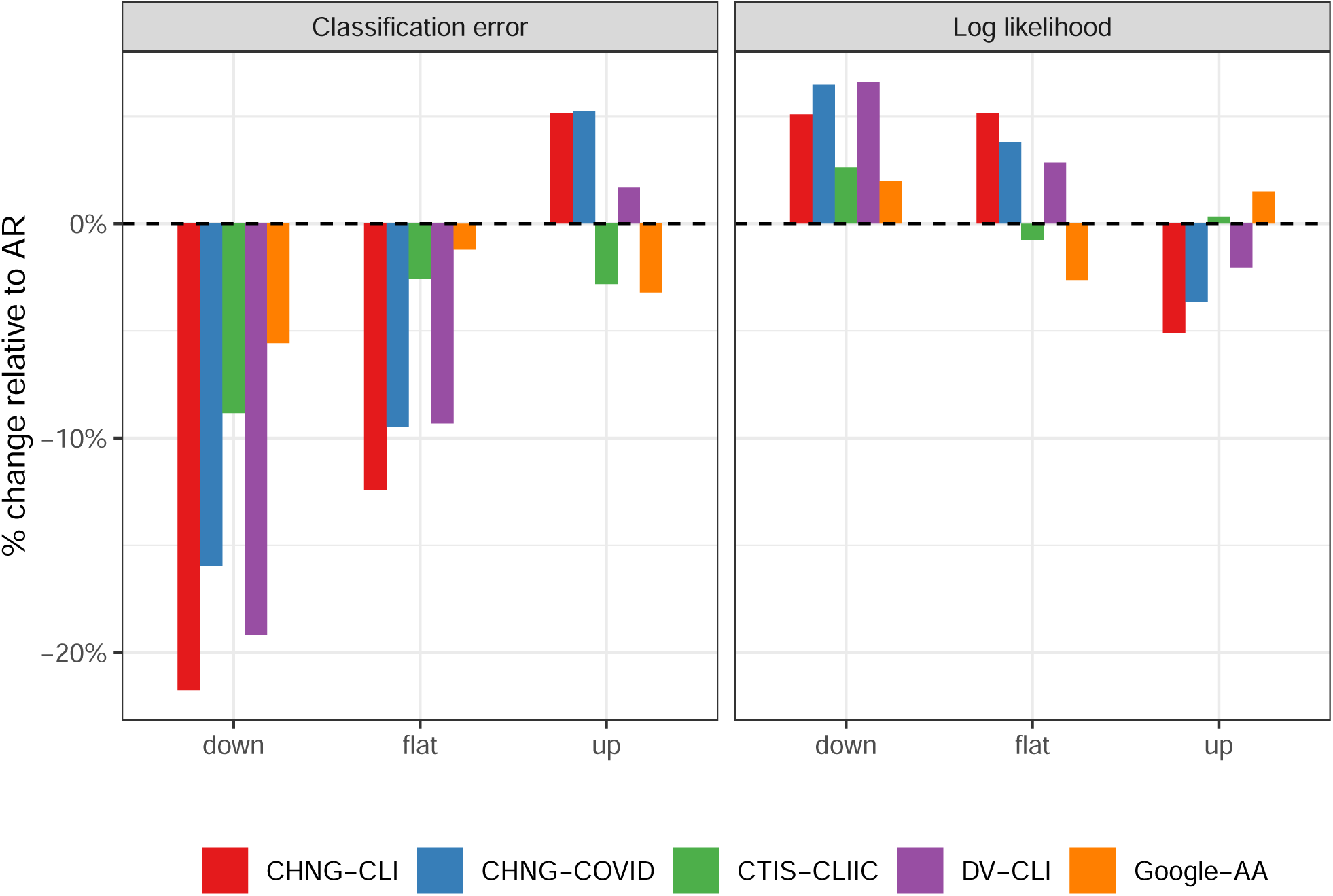
Percentage change in classification error and log likelihood, relative that of the AR model, separated into up, down, and flat periods. Like the analogous forecasting analysis, the indicator models generally do better during down and flat periods.

**Figure 19:**
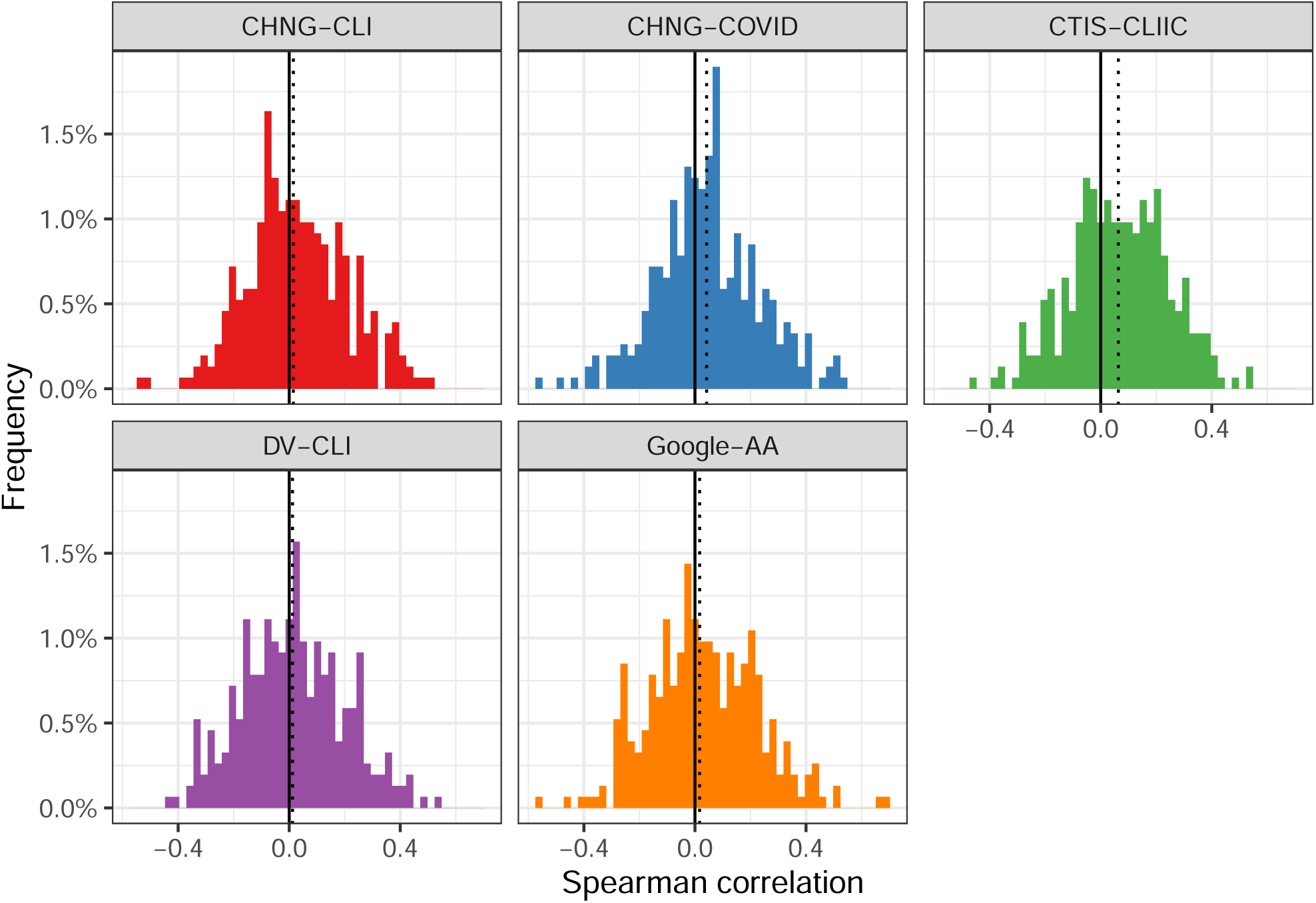
Histograms of the Spearman correlation between the ratio of forecaster WIS to AR WIS with the % change in case rates, relative to case rates 7 days earlier.

One hypothesis for diminished relative performance during upswings is that the AR model tends to overpredict downswings and underpredict upswings. Adding indicators appears to help avoid this behavior on the downswing but not as much on upswings. Figure 20 shows the correlation between between WIS(AR) −WIS(*F*) and the difference of their median forecasts. During downswings, this correlation is large, implying that improved relative performance of *F* is related to making lower forecasts than the AR model. The opposite is true during upswings. This is largely to be expected. However, the relationship attenuates in flat periods and during upswings. That is, when performance is better in those cases, it may be due to other factors than simply making predictions in the correct direction, for example, narrower confidence intervals.

**Figure 20:**
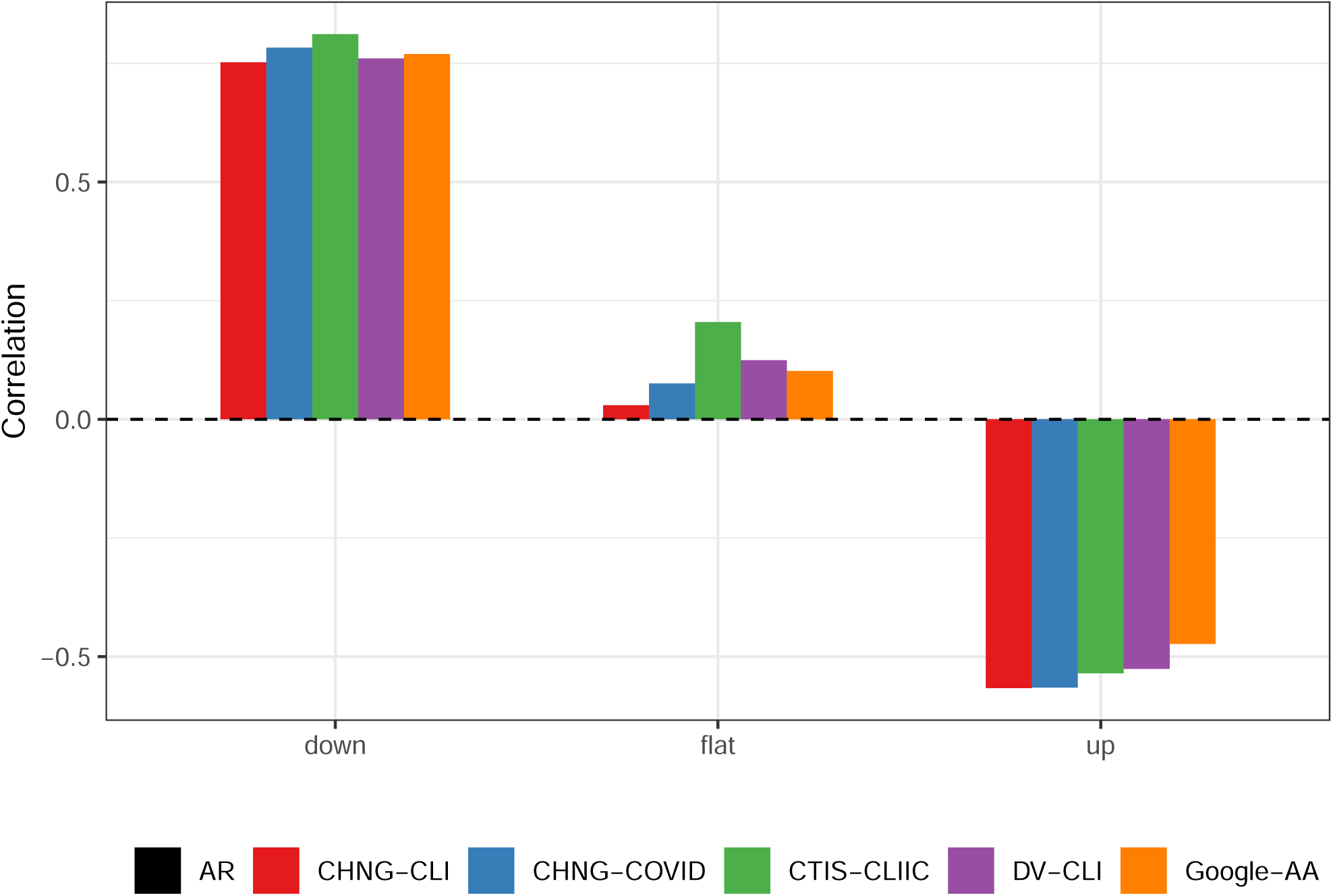
Correlation of the difference in WIS with the difference in median predictions (each difference being between the AR model and an indicator model), separated into up, down, and flat periods. In down periods, improvements in forecast error are highly correlated with lower median predictions. The opposite is true in up periods, but the conclusion here appears to be weaker.

It is important to note that, even though some indicators—notably CHNG-COVID and CHNG-CLI— underperform relative to the AR model during upswings, all models dramatically outperform the baseline in such periods. Furthermore, Figure 21 shows the performance of all forecasters relative to the baseline model. All forecasters suffer relative to the baseline during down periods, but the AR is the worst. In contrast, all models beat the baseline during up periods, even CHNG-COVID and CHNG-CLI, though not by quite as much as the AR does.

**Figure 21:**
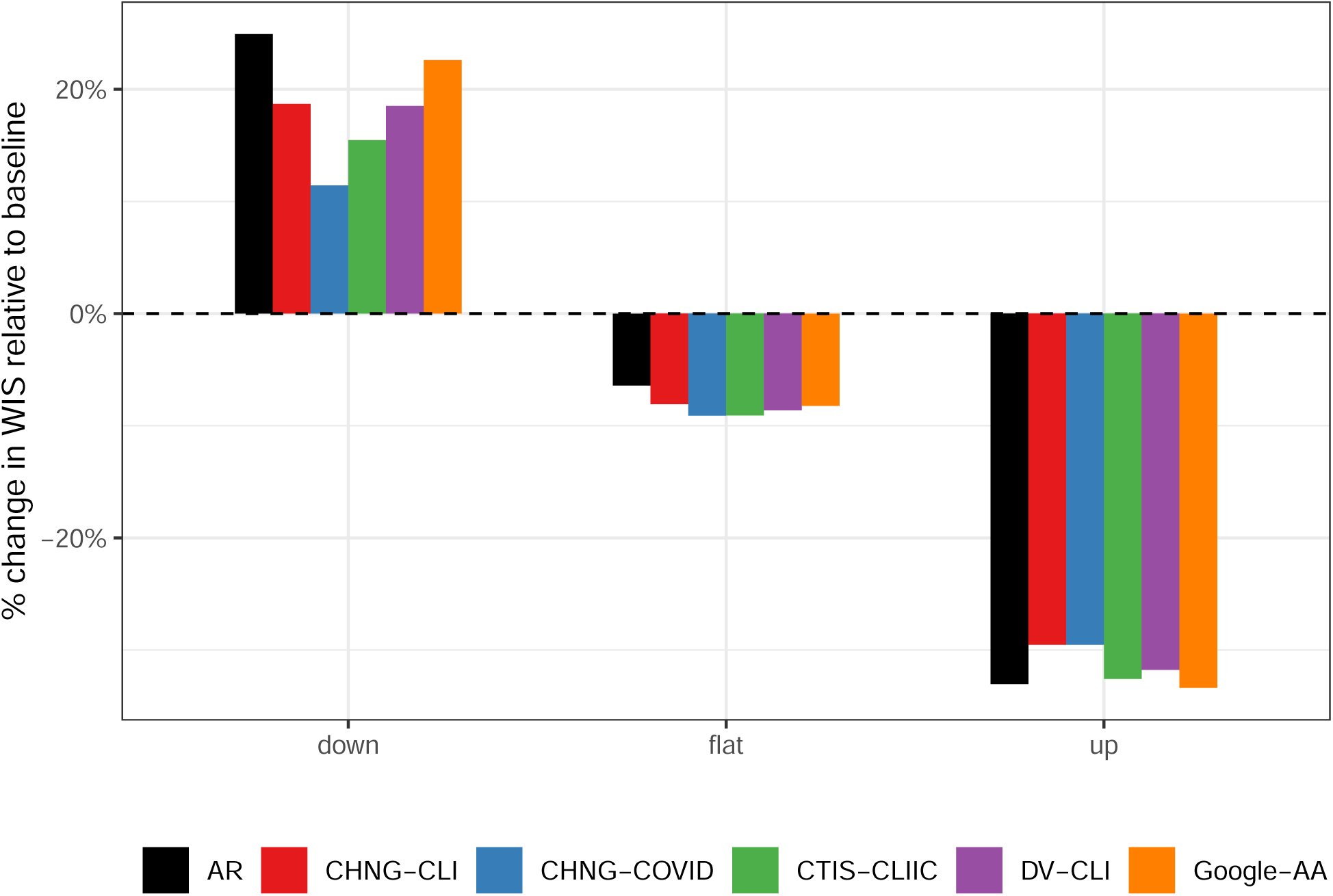
Percentage change in average WIS of the forecaster (AR or indicator assisted), relative to the baseline. All models perform poorly during down periods, but the indicators help. During flat periods, the indicators improve slightly over the AR. During up periods, all forecasters do much better than the baseline, but only some do as well as AR.

### G Leadingness and Laggingness

In Section 2.D of the main text, we discuss the extent to which the indicators are leading or lagging case rates during different periods. To define the amount of leadingness or laggingness at location *f*, we use the cross correlation function (CCF) between the two time series. The CCF_*ℓ*_(*a*) of an indicator series *X*_*ℓ*_ and case rate series *Y*_*ℓ*_ is defined as their Pearson correlation where *X*_*ℓ*_ has been aligned with the values of *Y*_*ℓ*_ that occurred *a* days earlier. Thus, for any *a >* 0, CCF_*ℓ*_(*a*) *>* 0 indicates that *Y*_*ℓ,t*_ is moving together with *X*_*ℓ,t*+*a*_. In this case we say that *X*_*ℓ*_ is lagging *Y*_*ℓ*_. For *a <* 0, CCF_*ℓ*_(*a*) *>* 0 means that *Y*_*ℓ,t*_ is positively correlated with *X*_*ℓ,t*−*a*_, so we say that *X*_*ℓ*_ leads *Y*_*ℓ*_.

Figure 22 shows the standardized signals for the HRR containing Charlotte, North Carolina, from August 1, 2020 until the end of September. These are the same signals shown in Figure 1 in the manuscript, but using finalized data. To define “leadingness” we compute CCF_*ℓ*_(*a*) (as implemented with the R function ccf()) for each *a* ∈ {−15, …, 15}using the 56 days leading up to the target date. This is the same amount of data used to train the forecasters: 21 days of training data, 21 days to get the response at *a* = 21, and 14 days for the longest lagged value. The orange dashed horizontal line represents the 95% significance threshold for correlations based on 56 observations. Any correlations larger in magnitude than this value are considered statistically significant under the null hypothesis of no relationship. We define leadingness to be the sum of the significant correlations that are leading (those above the dashed line with *a <* 0) while laggingness is the same but for *a >* 0. In the figure, there are three significant correlations on the “leading” side (at *a* = −5, −4, − 3), so leadingness will be the sum of those values while laggingness is 0: on September 28 in Charlotte, DV-CLI is leading cases leading but not lagging.

**Figure 22:**
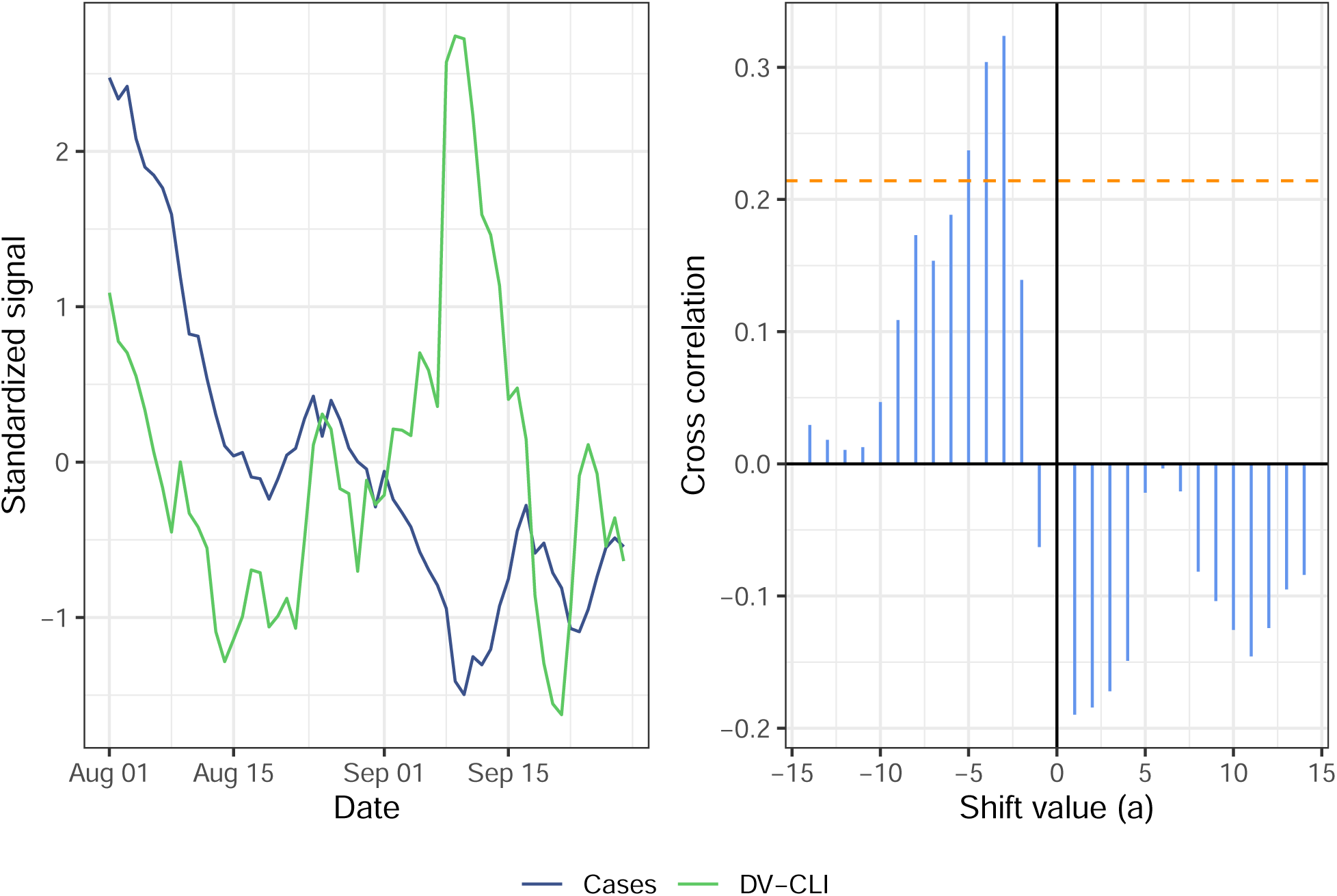
Illustration of the cross-correlation function between DV-CLI and cases. The left panel shows the standardized signals over the period from August 1 to September 28 (as of May 15, 2021). The right panel shows CCF_*ℓ*_(*a*) for different values of *a* as vertical blue bars. The orange dashed line indicates the 95% significance threshold. By our leadingness/laggingness metric, DV-CLI is leading (but not lagging) cases over this period.

Figure 23 shows the correlation between laggingness and the difference in forecaster WIS and AR WIS. Unlike leadingness (Figure 5 in the manuscript) there is no obvious relationship that holds consistently across indicators. This is encouraging as laggingness should not aid forecasting performance. On the other hand, if an indicator is more lagging than it is leading, this may suggest diminished performance. Figure 24 shows the correlation of the difference in leadingness and laggingness with the difference in WIS. The pattern here is largely similar to the pattern in leadingness described in the manuscript: the relationship is strongest in down periods and weakest in up periods with the strength diminishing as we move from down to flat to up for all indicators.

**Figure 23:**
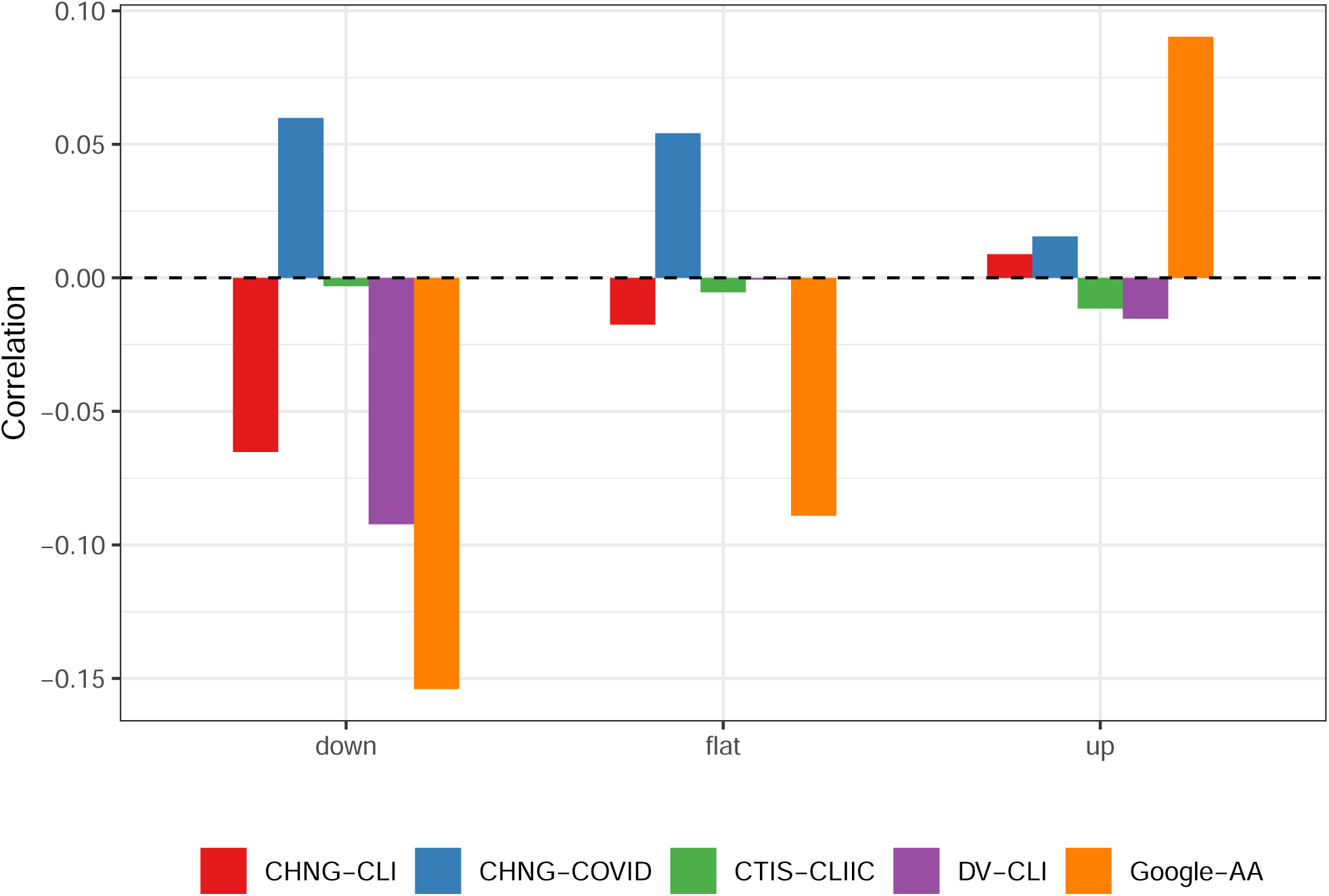
Correlation of the difference in WIS with the laggingness of the indicator at the target date, stratified by up, down, or flat period. Compare to Figure 5 in the manuscript.

**Figure 24:**
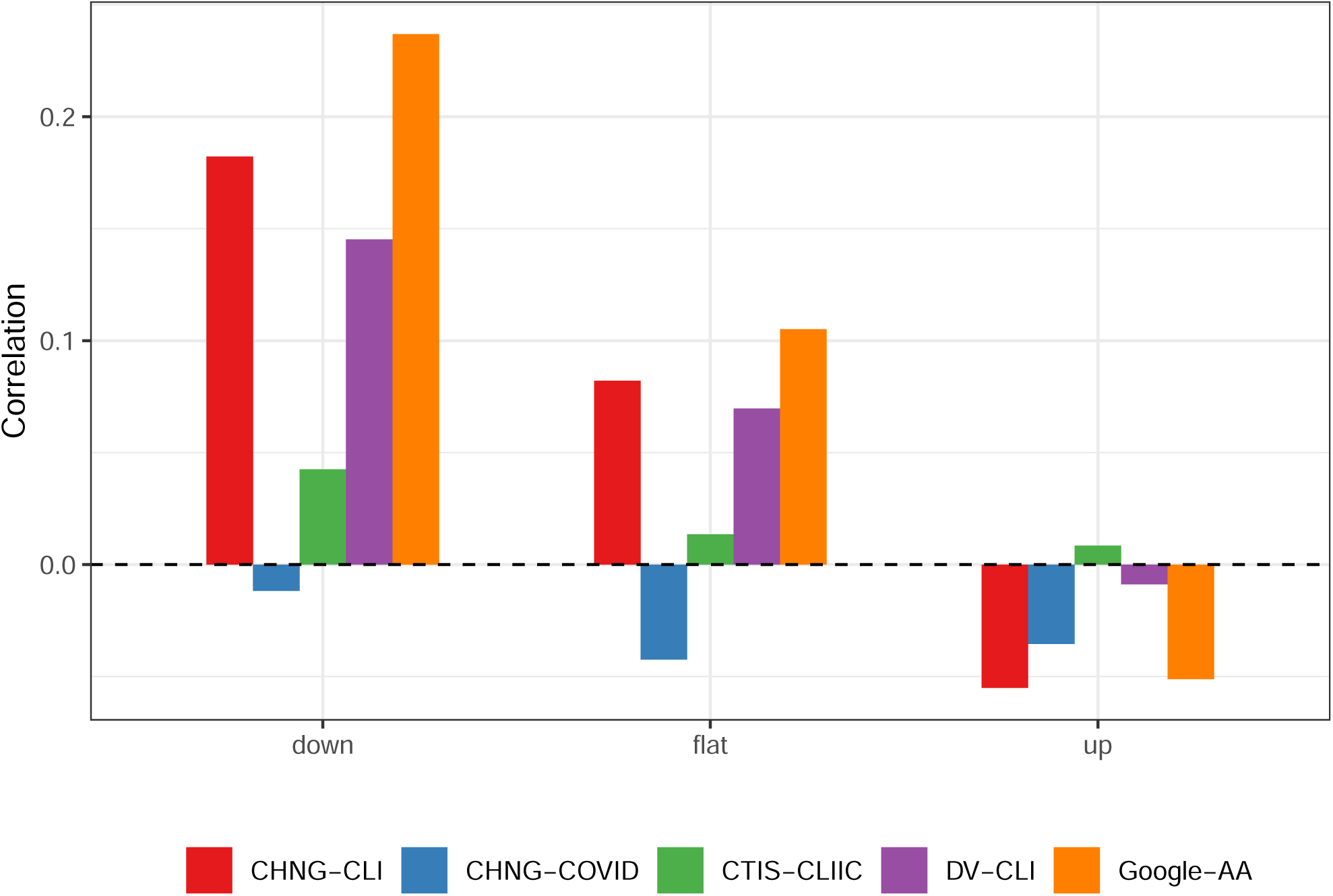
Correlation of the difference between leadingness and laggingness with the difference in WIS. The relationship is essentially the same as described in the manuscript and shown in Figure 5.

In calculating the CCF and the associated leadingness and laggingness scores, we have used the finalized data, and we look at the behavior at the target date of the forecast. That is we are using the same data to evaluate predictive accuracy as to determine leadingness and laggingness. It should be noted that the leadingness of the indicator at the time the model is trained may also be important. Thus, we could calculate separate leadingness and laggingness scores for the trained model and for the evaluation data and examine their combination in some way. However, we do not pursue this further.

### H Disaggregation Over Time and Space

Following the suggestion of an anonymous reviewer, we investigate two other disaggregated versions of the main forecasting result shown in Figure 3 on the manuscript. Below we use the term “error” to refer to the WIS summed over all ahead values. The first (Figure 25) displays the cumulative error, up through any point in time, of each forecaster divided by the cumulative error of the baseline. This perspective should illustrate how the models perform over time, drawing attention to any nonstationary behavior. During the initial increase in cases in July 2020, CTIS-CLIIC and Google-AA gain quite a bit acurracy compared to the AR model. All the indicators do much better than the AR model during the following downturn (the ebb of the second wave). The AR model actually improves over the indicators in October 2020, before losing a bit in late November.

**Figure 25:**
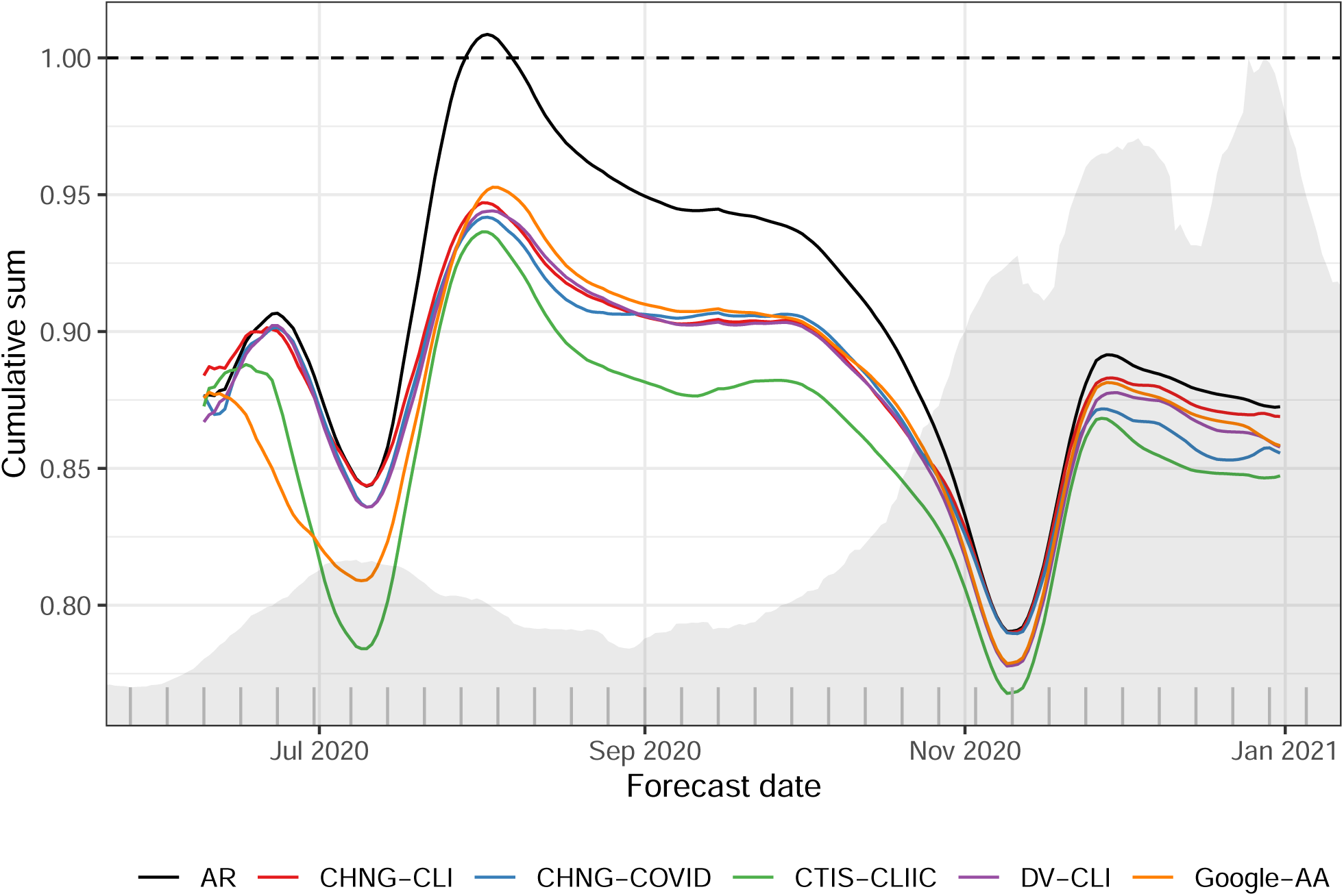
Cumulative sum of WIS for each forecaster divided by the cumulative sum of WIS for the baseline model. The shaded background shows national case incidence for the 14-day ahead target. Hash marks along the x-axis denote weeks.

Figure 26 examines the spatial behavior of errors over the entire period of the indicator models relative to the AR. For ease of comparison, we show the percent improvement in each HRR. Negative numbers (blue) mean that the indicator helped while positives (red) mean that the indicator hurt forecasting performance. The clear pattern is that in most HRRs, the indicators improved performance, though usually by small amounts (2.5%–10%). In some isolated HRRs, performance was markedly worse, though there does not appear to be any particular pattern to these locations. Interestingly, the geographic patterns of improvement differ quite a lot in between the indicators. This suggests that a forecasting model that carefully combines all of indicators could be a considerable improvement.

**Figure 26:**
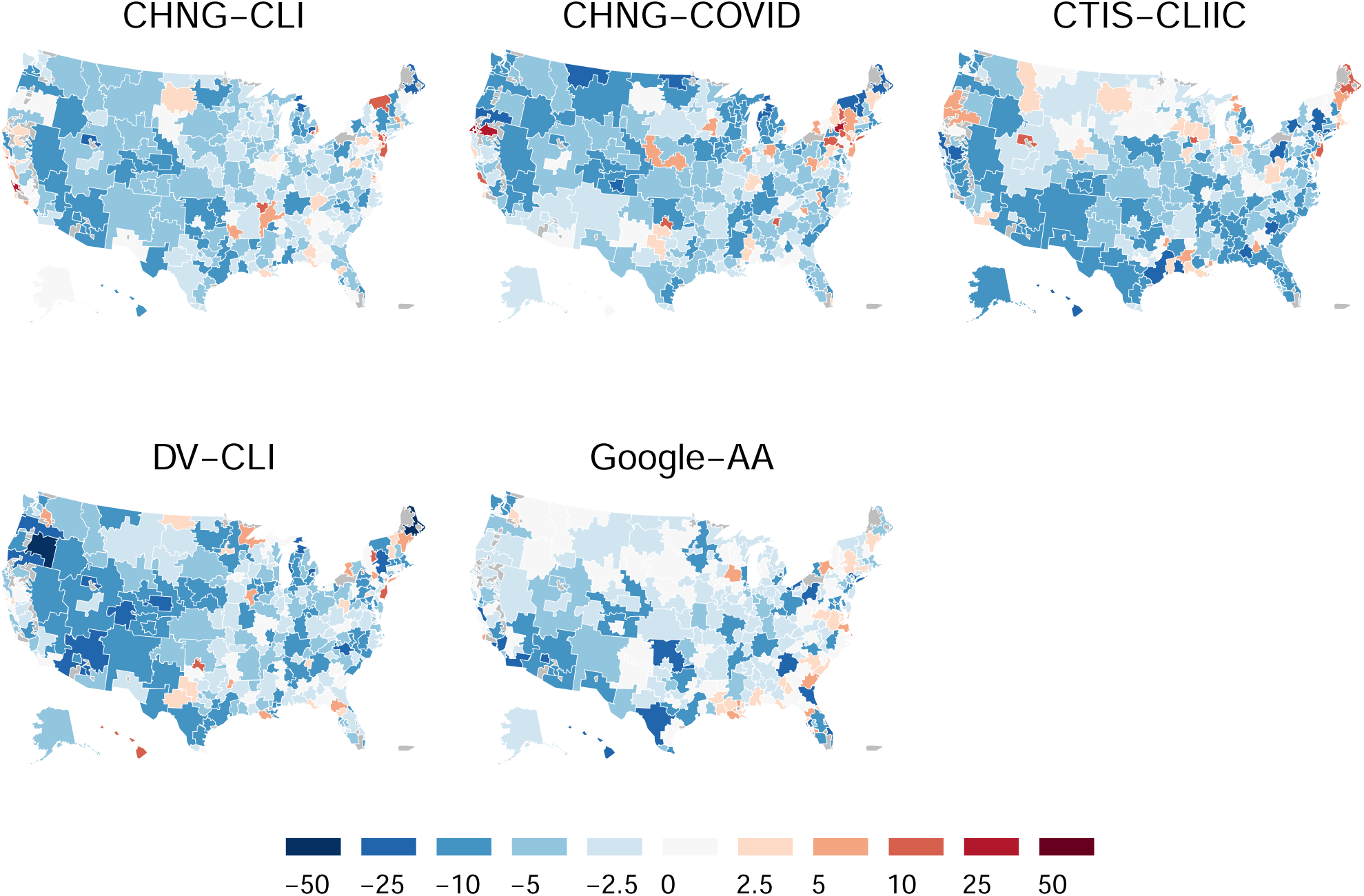
Percentage improvement in WIS, relative to the AR forecaster, by HRR (negative numbers indicate improved performance, positives indicate worsening).

### Footnotes

1 In a sense, this is implicitly estimating a nonparametric space-time effect for forecaster error, and assuming that has a shared, multiplicative contribution to forecaster errors. That is, if one imagines that a forecaster’s WIS is composed of multiplicative space-time effects *S*_,*t*_ shared across all forecasters, WIS(*F*_*ℓ,t,f*_, *Y*_*ℓ,t*_) = *S*_*ℓ,t*_*E*_*f,t*_ with *E*_*f,t*_ a forecaster-specific error, then taking the ratio of individual WIS values cancels these space-time effects.

